# Extracellular vesicle-bound DNA in urine is indicative of kidney allograft injury

**DOI:** 10.1101/2022.04.07.22273527

**Authors:** Ivana Sedej, Maja Štalekar, Magda Tušek Žnidarič, Katja Goričar, Nika Kojc, Polona Kogovšek, Vita Dolžan, Miha Arnol, Metka Lenassi

**Author notes:** **Correspondence:**Metka Lenassi, PhD, Institute of Biochemistry and Molecular Genetics, Faculty of Medicine, University of Ljubljana, Vrazov trg 2, 1000 Ljubljana, Slovenia, Miha Arnol, MD, PhD, Department of Nephrology, Division of Internal Medicine, University Medical Center Ljubljana, Zaloška cesta 7, 1000 Ljubljana, Slovenia. co-first authorship (Ivana Sedej, Maja Štalekar). co-senior authorship (Miha Arnol, Metka Lenassi). **Statements relating to our ethics and integrity policies:** The study was approved by the National Medical Ethics Committee of the Republic of Slovenia (approval Nr.: 0120-216/2019) and was conducted in accordance with the Declaration of Helsinki. An informed consent form was signed by all study participants. All mandatory laboratory health and safety procedures have been complied with in the course of conducting here reported experimental work. Maja Štalekar.

## Abstract

Extracellular vesicle-bound DNA (evDNA) is an understudied extracellular vesicle (EV) cargo, particularly in cancer-unrelated fundamental and biomarker research. Although evDNA has been detected in urine, little is known about its characteristics, localization, and biomarker potential for kidney pathologies. To address this, we enriched EVs from urine of well-characterized kidney transplant recipients undergoing allograft biopsy, characterized their evDNA and its association to allograft injury. Using DNase treatment and immunogold labelling TEM, we show that DNA is bound to the surface of urinary EVs. Although the urinary evDNA and cell-free DNA correlated in several characteristics, the DNA integrity index showed evDNA was less fragmented (*P* < 0.001). Urinary EVs from patients with rejection and non-rejection allograft injury were significantly larger (mean: *P* = 0.045, median: *P* = 0.031) and have bound more DNA as measured by normalized evDNA yield (*P* = 0.018) and evDNA copy number (*P* = 0.007), compared to patients with normal histology. Urinary evDNA characteristics associated with the degree of interstitial inflammation, combined glomerulitis and peritubular capillaritis, and inflammation in areas of fibrosis (all *P* < 0.050). The normalized dd-evDNA copy numbers differed between the antibody- and T cell-mediated rejection (*P* = 0.036). Our study supports the importance of DNA as urine EV cargo, especially as potential non-invasive kidney allograft injury biomarker.

## 1 Introduction

Liquid biopsy, a minimally invasive test performed on biofluid samples to detect distant pathology, is a promising alternative to the established invasive biopsies used in cancer and organ transplant diagnostics. Most studies focus on circulating cells, circulating RNA or cell-free DNA (cfDNA), with extracellular vesicles (EVs) emerging as a promising novel analyte (Szilágyi et al., 2020; Tamura, et al., 2021). EVs are a heterogeneous group of spherical membrane-bound particles, which play a prominent role in intercellular communication (Yáñez-Mó et al., 2015). They can differ in their biogenesis, cell of origin, size, molecular composition (nucleic acids, proteins, lipids, carbohydrates, and metabolites), and biological role (van Niel et al., 2018). All cells studied so far release EVs extracellularly, which *in vivo* accumulate to high concentrations in all body fluids. As stable carriers of (patho)physiological signals that reflect the state of the parental cell, they are being studied as biomarkers in various diseases (González & Falcón-Pérez, 2015). These studies focused mainly on miRNA, mRNA, and protein EV cargo, but neglected the EV-bound DNA (evDNA).

In blood, evDNA exists as single or double stranded molecule of genomic or mitochondrial origin, with size range from 200 bp in small EVs to up to >2 million bp in large EVs (reviewed in Malkin & Bratman, 2020). In cancer, evDNA was shown to represent the entire genome and mutational status of the cell of origin (Lázaro-Ibáñez et al., 2019; Vagner et al., 2018). DNA can be attached to EV surface or is present in its lumen (Lázaro-Ibáñez et al., 2019; Maire et al., 2021; Németh et al., 2017; Shelke et al., 2016; Vagner et al., 2018). Exact mechanism of DNA sorting is not known, but might include sequestering of cytosolic DNA during outward plasma membrane budding or intraluminal vesicles formation, or shuttling of collapsed micronuclei to multivesicular bodies (reviewed in Malkin & Bratman, 2020). The biological function of evDNA is poorly understood, but might include roles in cellular homeostasis, transfer of genetic material or immune response (Takahashi et al., 2017; Torralba et al., 2018). Despite the growing literature on evDNA in blood, there is a paucity of studies on DNA bound to urinary EVs (uEVs). Study by Lee et al. showed that urinary evDNA reflected tumor mutation status in nine urothelial carcinoma patients (Lee et al., 2018), but little is known about the evDNA characteristics, localization, and biomarker potential.

Kidney transplantation is a perfect model to study the biomarker potential of urinary evDNA. It is the most effective therapy for end-stage kidney disease, but unrecognized and subsequently untreated injury of the allograft can contribute to the loss of function over time and decreased allograft survival (Nankivell & Kuypers, 2011). Long-term allograft survival remains a major challenge, mostly due to acute and chronic rejection. Rejection can be broadly categorized as humoral, when mediated by the presence of anti-donor-specific antibodies (antibody-mediated rejection; ABMR), or cellular, when caused by infiltration of the interstitium by T cells and macrophages (T-cell mediated rejection; TCMR). Although the rate of acute rejection has decreased in the modern era of immunosuppression, recent reported incidence of acute rejections ranges from 11% to 26% (Christakoudi et al., 2019; Sigdel et al., 2018; Suthanthiran et al., 2013). This has been associated with poor long-term allograft survival (Opelz & Döhler, 2008). Serum creatinine, estimated glomerular filtration rate (eGFR), and urinary protein excretion are traditional biomarkers used to monitor the kidney allograft, but they lack sensitivity and specificity (Anglicheau et al., 2016). Histopathologic characterization of kidney biopsies based on the Banff classification remains the standard for diagnosis of kidney allograft injury (KDIGO Transplant Work Group, 2009; Solez et al., 1993). However, biopsies are invasive and costly procedures that can be associated with significant morbidity and are inappropriate for continuous monitoring (Hogan et al., 2016). In addition, the biopsy sample may not accurately represent the state of the entire kidney, and histopathologic interpretation is subject to inter-observer variability (Mengel et al., 2007). Therefore, novel non-invasive biomarkers are needed to allow frequent monitoring and earlier detection of kidney allograft injury.

Urinary EVs have a proven role in kidney physiology and are promoters of several kidney diseases (reviewed in Pomatto et al., 2017; Yates et al., 2022a, 2022b). They are released from cells of every nephron segment, bladder, and prostate in males, as well as resident immune cells, and are easily routinely accessible (Erdbrügger et al., 2021; Ranghino et al., 2015). As biomarkers of kidney allograft injury, elevated levels of uEV-bound tetraspanin-1, hemopexin (Lim et al., 2018) or specific set of mRNAs (El Fekih et al., 2021) were associated with rejection. Protein composition of uEVs also reflects delayed graft function, reduction in ischaemia-reperfusion injury, or the inflammatory and stress response of the kidney allograft (Alvarez et al., 2013; Braun et al., 2020; Oshikawa-Hori et al., 2019; Park et al., 2017). Studies have so far neglected uEV-bound DNA, although cell-free DNA (cfDNA) is a promising biomarker for allograft health and function (reviewed in Oellerich et al., 2021; Verhoeven et al., 2018). Organ transplantation is namely also genome transplantation, which allows specific detection of donor-derived cfDNA (dd-cfDNA) released from the damaged allograft into the recipient’s body fluids (Oellerich et al., 2021). The pivotal DART study has shown that dd-cfDNA levels > 1% in the plasma of recipients indicate kidney allograft rejection (Bloom et al., 2017). This has been confirmed by several larger cohort and multicenter studies, and absolute quantification of dd-cfDNA has been proposed as an alternative readout (Gielis et al., 2020; Halloran et al., 2022; Huang et al., 2019; Oellerich et al., 2019; Sigdel et al., 2018; Whitlam et al., 2019). In a rare study on urine, Sigdel et al. showed that urinary dd-cfDNA was significantly higher in patients with acute rejection than in patients with stable kidney allograft (Sigdel et al., 2013). Urinary cfDNA analysis is also part of the multimarker test for early detection of kidney allograft rejection (Watson et al., 2019; Yang et al., 2020). It would be interesting to evaluate the utility of urine evDNA as a biomarker of kidney allograft injury.

The aim of this study was to investigate DNA as a cargo of urinary EVs and to explore its utility as a biomarker for kidney allograft injury in patients undergoing surveillance or indication biopsy. To this end, we isolated EVs, evDNA, and cfDNA from the urine of well-characterized kidney transplant recipients, and characterized EV concentration and size by nanoparticle tracking analysis; and DNA yield, copy number, integrity index, and donor-derived DNA (ddDNA) by fluorometry, genotyping and droplet digital PCR (ddPCR). Localization of evDNA was investigated by DNase assay and immunogold labelling for electron microscopy. Finally, association of evDNA characteristics to patients’ clinical characteristics was tested to evaluate evDNA as a biomarker of kidney allograft injury.

## 2 Materials and Methods

### 2.1 Study design and data collection

Forty-one adult patients, who underwent a deceased donor kidney transplantation at the Center for Kidney Transplantation, Department of Nephrology, University Medical Center Ljubljana, Slovenia, were enrolled in this prospective observational study at the time of kidney allograft biopsy. From November 2018 to December 2020, 21 enrolled patients underwent surveillance biopsy 1 year after transplantation, whereas 20 patients underwent for-cause biopsy due to clinical indication (i.e., an increase in serum creatinine > 20% from baseline without apparent cause; a new-onset or increase in proteinuria). Exclusion criteria included multiple-solid organ transplants. Among the forty-one study participants, one patient was later confirmed to have active infection with BK polyomavirus and was excluded from further analysis. The study was approved by the National Medical Ethics Committee of the Republic of Slovenia (approval Nr.: 0120-216/2019) and was conducted in accordance with the Declaration of Helsinki and compliant with the Good Clinical Practice Guidelines. An informed consent form was signed by all study participants. All mandatory laboratory health and safety procedures were followed in performing the reported experimental work.

Fully anonymized patient data, collected at the time of biopsy, included patient *demographics* (age, sex), recipient and donor *clinical parameters* (expanded criteria donor (ECD; Kauffman et al., 1997), human leukocyte antigen (HLA) mismatch, delayed graft function (DGF), time from transplantation to biopsy, donor-specific antibodies (DSA)), and *laboratory data* (serum creatinine (S-creatinine), urine creatinine (U-creatinine), estimated glomerular filtration rate (eGFR), urinary estimated protein excretion rate (ePER) and serum C-reactive protein (S-CRP)). Histopathologic examination of blinded biopsy samples was performed at the Institute of Pathology (Faculty of Medicine, University of Ljubljana) by the dedicated pathologist, in accordance with the 2017 Banff Working Groups’ criteria (Haas et al., 2018). TCMR was reported as integrated tubulo-interstitial (borderline and grade IA/B) or vascular (grade IIA/B). The phenotypes of ABMR were classified as acute or chronic active. The diagnosis of acute ABMR was based on morphologic evidence of acute tissue injury (i.e., peritubular capillaritis and/or glomerulitis) and positive C4d staining. The diagnosis of chronic ABMR was based on the morphologic evidence of antibody-mediated chronic tissue injury, specifically glomerular double contours compatible with chronic glomerulopathy on light and/or electron microscopy. Biopsy specimen reports with a diagnosis of mixed ABMR and TCMR were grouped with the ABMR subgroup. All recipients were on an immunosuppressive regimen consisting of a calcineurin inhibitor, mycophenolate mofetil or azathioprine, and steroids. The study patient characteristics are presented in Table 1. Any missing patient data are clearly indicated.

**Table 1:**
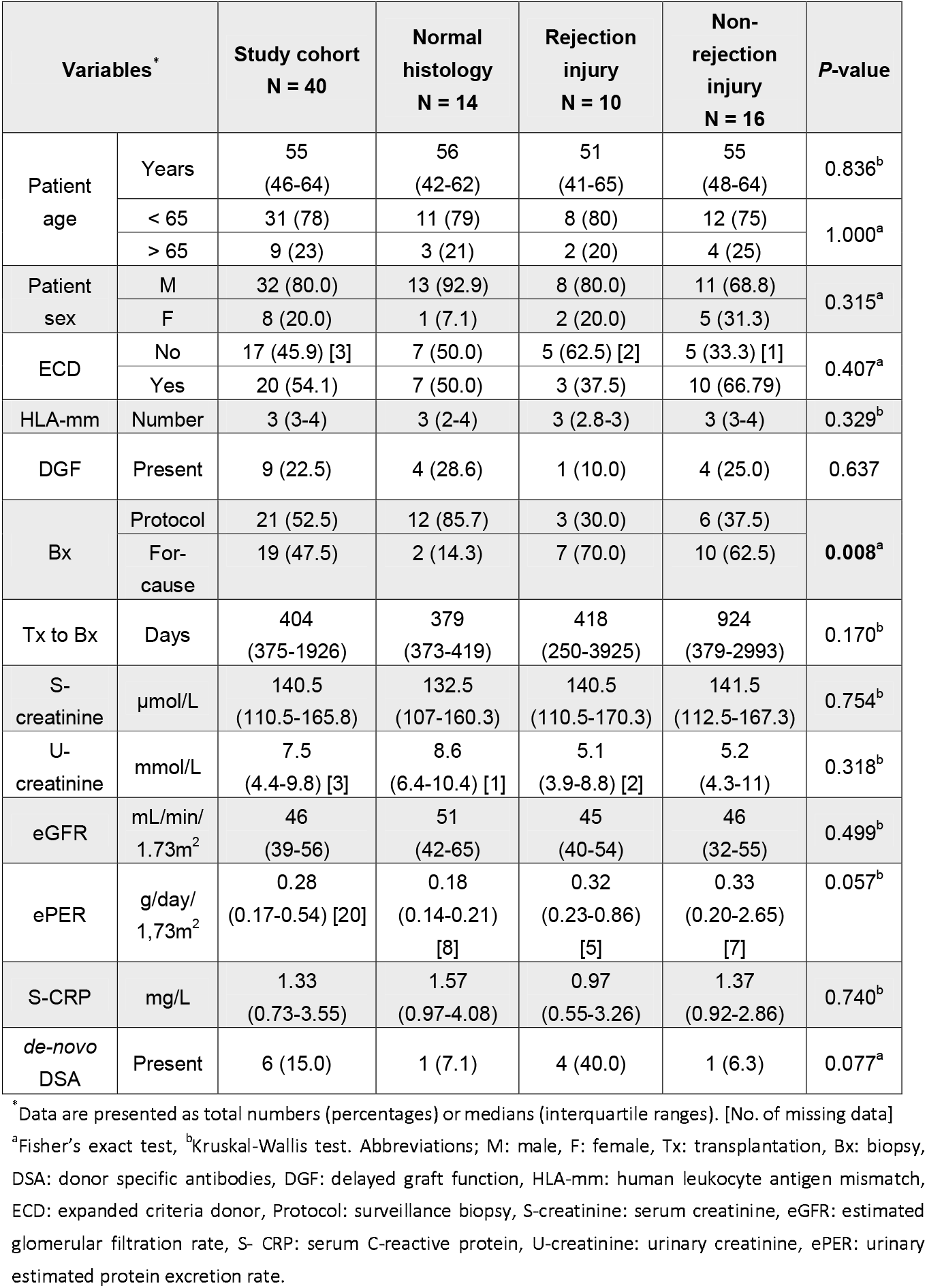
Kidney allograft recipients’ demographic, clinical and laboratory characteristics

Second morning spot urine and whole blood sampling was performed in the morning, prior to biopsy, and processed within four hours, always following the same protocols. *Peripheral whole venous blood* (4 mL) of patients was collected into ethylenediaminetetraacetic acid (EDTA) vacutainer tubes (BD Vacutainer, BD). Next, blood was centrifuged at 2,500 × g for 10 min at room temperature (RT). After removal of plasma, residual blood containing blood cells (2 mL) was stored at –20 °C for recipients’ genomic DNA extraction. *Second morning spot urine* of study patients was collected into sterile urine cups (Vacuette, Greiner Bio-one), transferred to conical polypropylene centrifuge tubes (TPP) and centrifuged at 2,000 × g for 15 min at RT. The supernatant of each patient’s urine sample was transferred to a fresh centrifuge tube, separated into two aliquots (for uEVs enrichment and cfDNA extraction) and stored at −80 °C. *Transplanted kidney tissue* was collected during the surveillance or for-cause kidney biopsy. Biopsy was performed by a medical professional, using the 16- or 18-gauge biopsy needle. A portion of one core kidney tissue sample (3 mm in length) was immediately stabilized in 500 µL of RNAlater Stabilization Solution (Thermo Fisher Scientific), incubated at RT for approximately 2 h to allow penetration into the tissue, and stored at – 20 °C for later donor genomic DNA extraction.

### 2.2 EV enrichment from urine

EVs were enriched from 20 mL of cell-free urine following our previously optimized protocol (Sedej et al., 2021). Briefly, upon urine thawing at RT, 400 µL of 0.5 M EDTA and 5 mL of 5X phosphate buffered saline (PBS, pH ∼ 7.4) were added for inhibition of uromodulin aggregation and Ca-oxalate formation, and pH neutralization, respectively (Kobayashi & Fukuoka, 2001). Samples were then concentrated to a volume of 500 µL on 100 kDa centrifugal filter units (Amicon, Merck Millipore), loaded on size-exclusion chromatography colons (SEC; qEVoriginal, Izon Q) for particle separation and eluted with 1X PBS. Twenty fractions of 500 µL were collected in protein low binding tubes (Eppendorf). Next, soluble protein content was determined by measuring absorbance at 280 nm (Synergy 2, BioTek) for each of the two combined fractions (1 mL). Protein-free SEC fractions 5–12 were pooled together and concentrated on 100 kDa centrifugal filter units (Amicon, Merck Millipore) to a volume of 70−80 µL, representing patient’s population of urine EVs (uEV sample). Samples were stored in DNA low binding tubes (Eppendorf) at −20 °C and processed further within one month.

### 2.3 Quantification of uEVs concentration and size

Nanoparticle Tracking Analysis (NTA) was used to determine the size and concentration of the enriched uEVs. Samples were diluted (400–3000X) in particle-free Dulbecco’s PBS (dPBS, Sigma-Aldrich) to reach an optimum concentration range of 1l×l10^7^–10^9^ particles/mL. Five 60-second movies per sample, at camera level 15, were recorded on the NanoSight NS300 instrument (488 nm blue laser) connected to the NanoSight Sample Assistant (Malvern Panalytical). Captured videos were manually examined and eliminated in the event of significant abnormalities, but not more than two videos per sample. Measurements were performed in duplicates. Raw data were analyzed by the NanoSight NTA 3.3 program at the following settings: water viscosity, temperature of 25 °C, detection threshold of 5, minimum track length of 10 and default minimum expected particle size and blur settings. Output data were expressed as uEV concentration, that is, the number of particles per mL of input urine, and uEV size, that is, the mean, modal, and median hydrodynamic diameter in nm.

### 2.4 DNase treatment of uEV samples

60 or 65 µL uEV samples were treated with 1 U of Ambion DNase I (Thermo Fisher Scientific) in 1X DNase buffer in 70 or 80 µL total reaction volume. Reactions were incubated at 37 °C for 30 min. After treatment, DNase was inactivated by incubation at 75 °C for 10 min or alternatively, denaturing buffer was added for inactivation and DNA extraction performed immediately.

To test the efficiency of DNase inactivation by heat, soybean genomic DNA was added to the uEV sample and incubated with either active DNase at 75 °C for 10 min or with heat-inactivated DNase at 37 °C for 30 min, to measure its activity during or after heat inactivation, respectively. To test DNase inactivation in denaturing buffer (buffer AL supplemented with proteinase K, QIAamp DNA Micro Kit), DNase was added to the soybean DNA-supplemented uEV sample, and denaturing buffer added immediately as the first step in the DNA extraction protocol. For all mentioned uEV samples, DNA extraction was performed and genomic DNA from soybean quantified by ddPCR.

### 2.5 DNA extraction

#### Genomic DNA extraction

For genotyping, donor genomic DNA was extracted from kidney tissue and recipient genomic DNA from study participants’ blood cells. For <u>donor genomic DNA</u> extraction, about 3 mm fragment of kidney tissue sample, stored in RNAlater solution (Thermo Fisher Scientific), was transferred to a fresh microcentrifuge tube (Eppendorf), and washed with 1X PBS for 15 min to remove recipient’s blood. Next, DNA was extracted using the QIAamp DNA Micro Kit (Qiagen), according to manufacturer’s instructions. In the last step, 20 µL of the elution buffer was applied to the center of the column membrane and incubated for 5 min at RT to increase the yield of extracted genomic DNA. After centrifugation, the elution step was repeated. DNA concentration was determined by measuring the absorbance at 260 nm (Synergy 2, BioTek) and the samples frozen at –20 °C.

For <u>recipient genomic DNA</u> extraction, the residual blood cells (2 mL) were thawed at RT and the recipient’s genomic DNA extracted with the EZNA SQ Blood DNA Kit II (Omega Bio-tek) as per manufacturer’s protocol for 1–3 mL Whole Blood. After pelleting the cells (protocol step 4), 10 mL of distilled water was added, and the centrifugation step repeated. After discarding the supernatant, the DNA precipitate was air dried. Genomic DNA was dissolved in 500 µL elution buffer in a water bath heated to 65 °C for 2 hours. The concentration of extracted DNA was determined by measuring the absorbance at 260 nm (Synergy 2, BioTek) and the sample frozen at –20 °C.

#### Cell free DNA and EV-bound DNA extraction

To optimize <u>cfDNA</u> extraction, 4 mL of pooled urine samples were processed (in duplicate) with five different kits according to the manufacturer’s protocols: MagMAX Cell-Free DNA Isolation Kit (Applied Biosystems), Apostle MiniMax High Efficiency cfDNA Isolation Kit (Beckman Coulter), NextPrep-Mag Urine cfDNA Isolation Kit (Perkin Elmer), Quick-DNA Urine Kit (Zymo Research), and QIAamp Circulating Nucleic Acid Kit (Qiagen). Next, cfDNA extractions were repeated on another urine pool with the three best performing kits. All cfDNA extractions from the urine of patients included in the cohort were performed using the Quick-DNA Urine Kit, and the cfDNA eluted with 15 µL of elution buffer.

<u>evDNA</u> was extracted with QIAamp DNA Micro Kit (Qiagen) following the manufacturer’s protocol for extraction of DNA from small volumes of blood. Use of carrier RNA was omitted. Elution of DNA was performed twice, each time adding 20 µL of elution buffer and incubating 5 min at RT before centrifugation.

cfDNA and evDNA concentrations were measured using Qubit dsDNA HS Assay Kit and Qubit 3 Fluorometer (both Invitrogen). Determined DNA concentration was used to calculate *DNA yield*.

### 2.6 Primers and probes

Custom TaqMan SNP Genotyping Assays (Thermo Fisher) used for genotyping of single nucleotide polymorphisms (SNPs; rs1707473, rs2691527, rs7687645, rs1420530, rs9289628, rs6070149) and quantification of ddDNA were described and validated previously on blood plasma (Jerič Kokelj et al., 2021). SNPs were chosen based on their potential to distinguish DNA of two individuals and are characterized with high minor allele frequency, low global fixation index, proximity to cfDNA fragment peaks and no overlap with repetitive elements. Amplicon lengths of SNP assays were 93, 70, 95, 67, 85 and 75 bp, respectively. RPP30 (amplicon length of 62 bp) and RPPH1 (amplicon length of 135 bp) assays were described previously (Oscorbin et al., 2019; Shebanits et al., 2019). RPPH1 and RPP30 were selected because they are single copy genes per haploid human genome and are commonly used as reference genes in copy number variation studies (Imaizumi et al., 2019; Oscorbin et al., 2019; Shebanits et al., 2019). SRY assay for quantification of presence of chromosome Y was designed as follows: forward primer 5’-CGAAGTGCAACTGGACAACAG-3’, reverse primer 5’-TAGCTGGTGCTCCATTCTTGAG-3’ and probe 5’-ACAGGGATGACTGTACGAAAGCCACACA-3’ (HEX, ZEN-3IABkFQ) (Integrated DNA Technologies), with amplicon length of 76 bp. Soybean Le1 assay (Eurofins) was described previously (De Broe et al., 1987).

### 2.7 Genotyping real-time PCR

Extracted genomic DNA from blood (recipient) and kidney tissue (donor) was genotyped by real-time PCR for six SNP loci using Custom TaqMan SNP Genotyping Assays (Thermo Fisher). 5 µL reactions with TaqMan Genotyping Master Mix (Applied Biosystems) and 5–10 ng genomic DNA per reaction were run in a 384-well format on the ViiA 7 Real-Time PCR System (Applied Biosystems) using the following thermal profile: 60 °C for 30 s (DC), 95 °C for 10 min, 40 cycles of 95 °C for 15 s and 60 °C for 1 min 30 s (DC), and finally 60 °C for 30 s (DC). The ramp rate was set to 1.6 °C/s for all steps. Data were collected in the steps marked with (DC). Positive controls prepared with gBlock Gene Fragments (Integrated DNA Technologies; described in detail in Suppl. material) or genomic DNA with known genotype were included for all genotypes. When donor genotype was not clear, due to contamination of donor kidney tissue with recipient’s blood, donor genomic DNA was additionally genotyped by ddPCR by comparing copy numbers of the six SNP amplicons and/or the SRY amplicon in allografts that were sex-mismatched, and subtracting the background copy number of known patient’s genotype.

### 2.8 Preamplification of DNA

Targeted multiplex PCR preamplification of six SNP loci, RPP30 and RPPH1 was performed on cfDNA and evDNA samples included in the study. As previously described (Andersson et al., 2015), 100 µl PCR reactions were prepared with 40 nM final concentration of each of 16 primers using Q5 Hot Start High-Fidelity 2X Master Mix (New England BioLabs). Denaturation at 98 °C for 3 min was followed by 11 or 12 amplification cycles (cfDNA and evDNA, respectively) of 10 seconds at 98 °C, 3 minutes at 64 °C and 20 seconds at 72 °C, and a 2 min final elongation step at 72 °C. When finished, reactions were diluted 10X in Tris EDTA and either analyzed with ddPCR in the same day or stored at –20 °C until analysis.

For validation of preamplification reaction, DNA mixtures of either gBlocks or genomic DNA with known genotypes were prepared (described in detail in Suppl. material). Next, the DNA copy numbers of the six SNPs, RPP30 and RPPH1 were determined by ddPCR in each sample, and values compared between samples without or with preamplification. Preamplification efficiency was determined as the ratio between measured copy number after preamplification and theoretically expected copy number after preamplification. Potential bias of preamplification reaction was assessed by comparing fractions of SNP genotypes, determined in the same manner as the *ddDNA fraction*, in the prepared DNA mixtures without and with preamplification. To test potential inhibitory effect of possible chemical contaminants in DNA eluates, the mock extractions of cfDNA or evDNA (extraction from buffer instead of biological samples) were added to preamplification reactions (described in detail in Suppl. material).

### 2.9 Droplet digital PCR

QX100 and QX200 Droplet Digital PCR Systems (Bio-Rad) were used. Reactions were prepared using ddPCR Supermix for Probes (No dUTP) (Bio-Rad) and up to 9 µL of preamplified cfDNA or evDNA. In cases of higher yields of DNA, the preamplified DNA was additionally diluted with nuclease-free water to avoid saturation of the ddPCR reaction. For quantification of RPP30 and RPPH1 amplicons, duplex ddPCR reactions were prepared using final concentration of 900 nM primers and 250 nM probes (all from Integrated DNA Technologies). The same six SNP assays were used for quantification of ddDNA as for genotyping of genomic DNA at 1X concentration. The SRY assay was performed in duplex format along with RPP30, as described for RPP30 and RPPH1 quantification. Soybean DNA was quantified using final concentration of 650 nM primers and 180 nM probe for Le1 (Eurofins). All reactions were performed in triplicate. Droplets were generated using the QX100, QX200, or AutoDG droplet generator (Bio-Rad). PCR cycling conditions were 95 °C for 10 min, 40 cycles of 94 °C for 30 s, and 60 °C for 1 min 30 s, with a final incubation step of 98 °C for 10 min. The ramp rate was set at 1.5 °C/s. Droplets were read using the QX100 or QX200 reader (Bio-Rad). QuantaSoft software version 1.7.4 (Bio-Rad) was used to manually determine the thresholds. The raw data were then exported and analyzed in Excel (Microsoft).

The initial copy number of an amplicon (before preamplification) was calculated as the amplicon copy number determined by ddPCR, adjusted for the dilution factor, and divided by the theoretical factor of PCR preamplification assuming 100% efficiency (2^11^ in the case of 11 and 2^12^ in the case of 12 cycles, respectively). Samples with initial copy number less than 5 were designated as insufficient DNA copy number and excluded from further analysis. An exception was made for RPP30 quantification of DNase-treated EV samples to allow analysis of samples after DNA degradation, but those samples were flagged as samples with less reliable quantification. The *DNA copy number* refers to the average of initial RPP30 copy numbers calculated from three separate measurements by ddPCR.

The *ddDNA fraction* in the samples was calculated from the data obtained by the SNP ddPCR reactions. For each sample, at least two informative SNP assays were identified that allowed us to distinguish between recipient and donor DNA. When recipients had a homozygous genotype (AA) and donors had a heterologous homozygous (BB) or heterozygous (AB) genotype, ddDNA fractions were calculated as in Beck et al. (2018), 100 × B / (A + B) for a homozygous donor and 2 × 100 × B / (A + B) for a heterozygous donor. When recipients had a heterozygous genotype (AB) and donors had a homozygous genotype (AA), ddDNA fractions were calculated as 100 × (A – B) / (A + B). The average ddDNA fraction, standard deviation, and coefficient of variation for each sample were calculated from data of at least two SNP assays, each analyzed in triplicate. The *ddDNA copy number* was calculated as the ddDNA fraction multiplied by the DNA copy number.

*DNA integrity index* was determined by calculating the ratio of RPPH1 (135 bp) to RPP30 (62 bp) copy numbers, representing the ratio of large to short amplicons (Mouliere et al., 2011).

DNA yield, DNA copy number and ddDNA copy number were normalized to U-creatinine, which was measured in mmol/mL.

### 2.10 Electron microscopy analysis of uEVs

To visualize uEVs, transmission electron microscopy (TEM) was carried out. Briefly, upon defrosting, 4 µL of uEV samples were adsorbed for 3 min at RT on freshly glow discharged, formvar coated and carbon stabilized copper grids (SPI), and stained with 1% (w/v) water solution of uranyl-acetate (SPI).

For detection of dsDNA in uEV samples, negative staining method was combined with immunogold labelling at electron microscopy level. 3 µL of defrosted uEV samples were applied on freshly glow discharged copper grids for 3 minutes, followed by floating the grids on a drop of 1% bovine serum albumin (Sigma) in 0.1 M phosphate buffer to block non-specific antibody binding, and then incubated on drop of mouse monoclonal antibodies against dsDNA (Abcam, ab 27156). After washing the grids, we incubated them on the drop of goat-anti-mouse IgG (Aurion), labelled with gold particles with 6 nm in diameter (Aurion Anionic Gold Tracers). Finally, the grids were stained with 1% (w/v) water solution of uranyl-acetate. As a negative control, anti-dsDNA antibodies were omitted from the labelling procedure for each uEV sample. Additionally, all uEV samples were also analyzed only by negative staining method.

The grids were observed by transmission electron microscope TALOS L120 (Thermo Fisher Scientific), operating at 100 kV. At least 10 grid squares were examined thoroughly, and representative micrographs (camera Ceta 16M) were taken at different places on the grid. The Velox software (Thermo Fisher Scientific) was used to process images.

### 2.11 Statistical analysis

Statistical analysis was performed using IBM SPSS Statistics, version 27.0 (IBM Corporation, Armonk, NY, USA). Continuous variables were described as median and interquartile range (25–75%), whereas categorical variables were described using frequencies. Fisher’s exact test was used to compare the distribution of categorical variables between the different groups. Non-parametric Kruskal-Wallis and Mann-Whitney tests with Benjamini-Hochberg adjustment for pairwise comparisons were used to compare the distribution of continuous variables between the different groups. The Wilcoxon signed rank test was used for comparison of related samples. Spearman’s rho correlation coefficient (ρ) was used to assess correlations between continuous variables. All statistical tests were two sided and the significance level was set at 0.05.

Figures were prepared in GraphPad Prism, version 9.3.1 (for Mac OS X, GraphPad Software), RStudio, version 1.4.1106 (RStudio Team, 2020), using packages ggplot2, version 3.3.5 (Wickham, 2016), ggpubr, version 0.4.0, finalfit, version 1.0.3, gridExtra, version 2.3 and cowplot, version 1.1.1. In the boxplots, the median, first and third quartiles are presented. Whiskers extend up/down to the largest/smallest value no further than 1.5 × inter-quartile range from the hinges. Data beyond the end of the whiskers were defined as outliers.

### 2.12 Data availability

We have submitted all relevant experimental data to the EV-TRACK knowledgebase (EV-TRACK ID: EV210292) (Van Deun et al., 2017).

## 3 Results

### 3.1 SEC-based method enriches pure EVs from the urine of kidney allograft recipients

To investigate DNA as uEV cargo, we first enriched EVs from urine of 40 enrolled patients, following a previously established protocol based on SEC (Figure 1A; Sedej et al., 2021). Because the phenotype of kidney allograft injury could influence the presence of DNA in uEV samples, study patients were divided into three groups according to histological assessment of the allograft. TCMR (N = 6) and acute and chronic active ABMR (N = 4) were integrated to form the rejection group (N = 10), and we distinguished them from samples with no major abnormalities (normal histology group; N = 14), and samples that were classified as having non-rejection injury (non-rejection group; N = 16). Non-rejection injury included specimens with isolated Banff scores (interstitial inflammation (i), tubulitis (t), and interstitial inflammation in the areas of fibrosis (i-IFTA)) that did not meet rejection criteria, calcineurin inhibitor nephrotoxicity, or recurrence of original kidney disease. The three patient groups did not differ in most of the characteristics studied, except in proportion of for-cause biopsies (*P* = 0.008) that was higher in the rejection injury group (Table 1). The median time (interquartile range) between transplantation and biopsy was 1943 (378–3646) days for for-cause biopsies and 382 (373–412) days for surveillance biopsies (*P* = 0.005).

**Figure 1:**
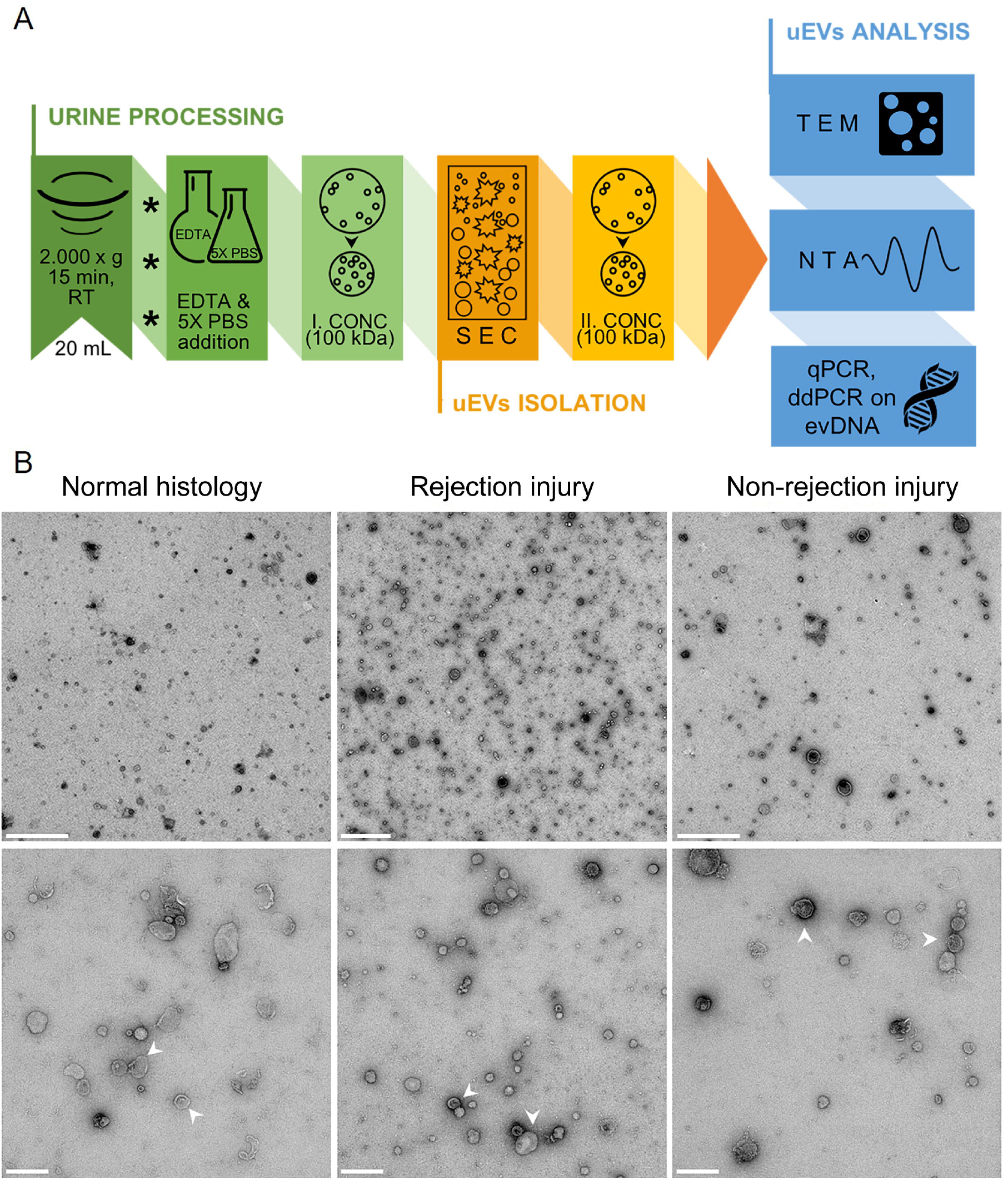
Size exclusion chromatography-based method enriches pure extracellular vesicles from urine of kidney allograft recipients. (A) Enrichment of EVs from urine (uEV): the processing of second morning spot urine upon collection is presented in green (asterisks indicate sample freezing at −80 °C before further processing), the uEV isolation step in orange, and the uEV analyses methods in blue. See Materials and Methods section for details. (B) Transmission electron micrographs of representative negative stained uEV samples from the normal histology (left), rejection injury (middle) and non-rejection injury (right) patient groups. White arrows indicate two (among many) uEVs. Scale bars: 1 µm (top row), 200 nm (bottom row). Abbreviations; RT: room temperature, EDTA: ethylenediaminetetraacetic acid, PBS: phosphate buffered saline, CONC: concentration, SEC: size exclusion chromatography, TEM, transmission electron microscopy, NTA, nanoparticle tracking analysis, qPCR, quantitative real-time PCR, ddPCR, digital droplet PCR, evDNA: extracellular vesicle-bound DNA.

To determine the purity of the uEV samples, representative uEV samples from the three patient groups were analyzed by TEM (Figure 1B). Representative micrographs show that the SEC-based method efficiently enriched EVs, with typical morphology, from the urine of all patient groups. The uEV samples were in general free of impurities, with visible proteins present in the sample from the patient with rejection injury (Figure 1B middle). At the same time, this sample displayed higher number of small EVs (less than 100 nm) compared to the uEV samples from patients with normal histology or with non-rejection injury. To summarize, using the SEC-based protocol we were able to enrich pure EVs from the urine of kidney transplant recipients, regardless of the phenotype of allograft injury.

### 3.2 Urine EVs size reflects the kidney allograft injury

Next, concentration and size of patients’ uEVs were determined by NTA. To account for inter-patient variability in overall urine concentration, uEV concentration values were normalized to U-creatinine, previously identified as a reliable normalization marker (Blijdorp et al., 2021). For three of the patients, the U-creatinine data was not available, so these samples were excluded from further analysis. The median (interquartile range) normalized uEV concentration in the patient cohort was 8.47 (3.63–15.10) × 10^10^/mmol U-creatinine, whereas the patients’ uEV mean, mode and median diameter were 169.9 nm (147.8–183.7), 125.8 nm (112.6–144.6), and 153.1 nm (133.8–172.0), respectively (Figure 2A, C, E, G). The NTA size distribution profiles of uEVs for each included patient are presented in Suppl. Figure 1. When comparing the normalized uEV concentration and uEV size among the patient groups (Figure 2), the normalized uEV concentrations were slightly higher in patients with rejection and non-rejection injury than in those with normal histology. However, the difference was not statistically significant (*P* = 0.626; Figure 2B). On the other hand, the uEV mean (*P* = 0.045) or median (*P* = 0.031) diameter were significantly higher in the rejection and non-rejection injury group compared with the normal histology group (Figure 2D, F, H). Overall, the uEV characteristics of patients with rejection and non-rejection injury differed compared to patients with normal histology.

**Figure 2:**
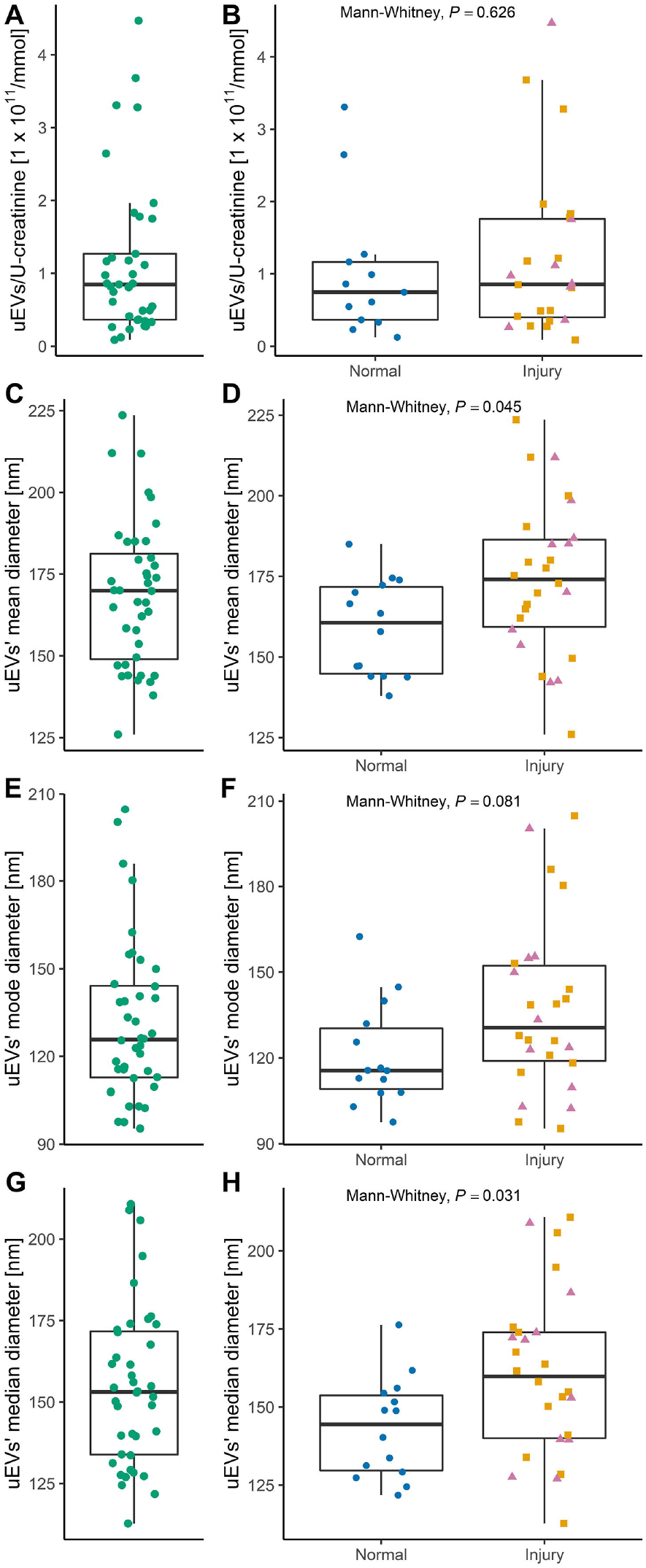
uEV characteristics of kidney allograft recipients as measured by Nanoparticle Tracking Analysis. Distribution of uEV concentrations [particles/mL urine] normalized to U-creatinine [mmol/mL] in (A) the patient cohort (N = 37) and in (B) patients with normal histology (N = 13) and with rejection (N = 8) or non-rejection (N = 16) injury. Distribution of mean (C, D), mode (E, F), or median (G, H) diameters of uEVs in the patient cohort (N = 40) and in patients with normal histology (N = 14) and with rejection (N = 10) or non-rejection (N = 16) injury. Data of patients with normal histology are presented as blue circles, rejection injury as purple triangles and non-rejection injury as orange squares. Mann-Whitney test was used to compare the uEV characteristics between patient groups.

### 3.3 Quantity of urine EV-bound DNA reflects the kidney allograft injury

To investigate if DNA is a cargo of EVs in urine, DNA was extracted from patients’ uEV samples. In parallel, cfDNA was extracted directly from the same urine samples for comparison. To improve efficiency of cfDNA extraction from urine, five commercially available kits were tested using the same pooled urine. A comparison of cfDNA yield and other characteristics for each kit are described in more detail in Supplementary Material (Suppl. Table 1, Suppl. Figure 2). Quick-DNA Urine Kit (Zymo Research) was selected for further work, because it gave the highest and most consistent yields, and its protocol is less laborious compared to other kits. Next, cfDNA was extracted from freshly thawed urine collected from all study patients.

The yield of extracted evDNA and cfDNA varied considerably, with a median of 3.4 ng (2.0–6.6) for evDNA and 18.5 ng (8.4–53.0) for cfDNA (Figure 3A). Nevertheless, the yields of evDNA and cfDNA correlated well (ρ = 0.680, *P* < 0.001, Figure 3B). To compensate for the differences in the overall concentration status of the urine samples, the DNA yields were normalized to U-creatinine (Figure 3C). Samples from the three patients for whom the U-creatinine data were not available were excluded from further analysis. The normalized DNA yields in the three groups were statistically significantly different *(P =* 0.042 for evDNA and *P* = 0.013 for cfDNA) (Figure 3D). More urinary evDNA and cfDNA were obtained in patients with rejection injury (*P* = 0.030 for evDNA and *P* = 0.018 for cfDNA) and non-rejection injury (*P* = 0.033 for cfDNA), than in patients with normal histology. Significance was maintained even when comparing the rejection and non-rejection injury combined with the normal histology group (*P* = 0.018 for evDNA and *P* = 0.004 for cfDNA).

**Figure 3:**
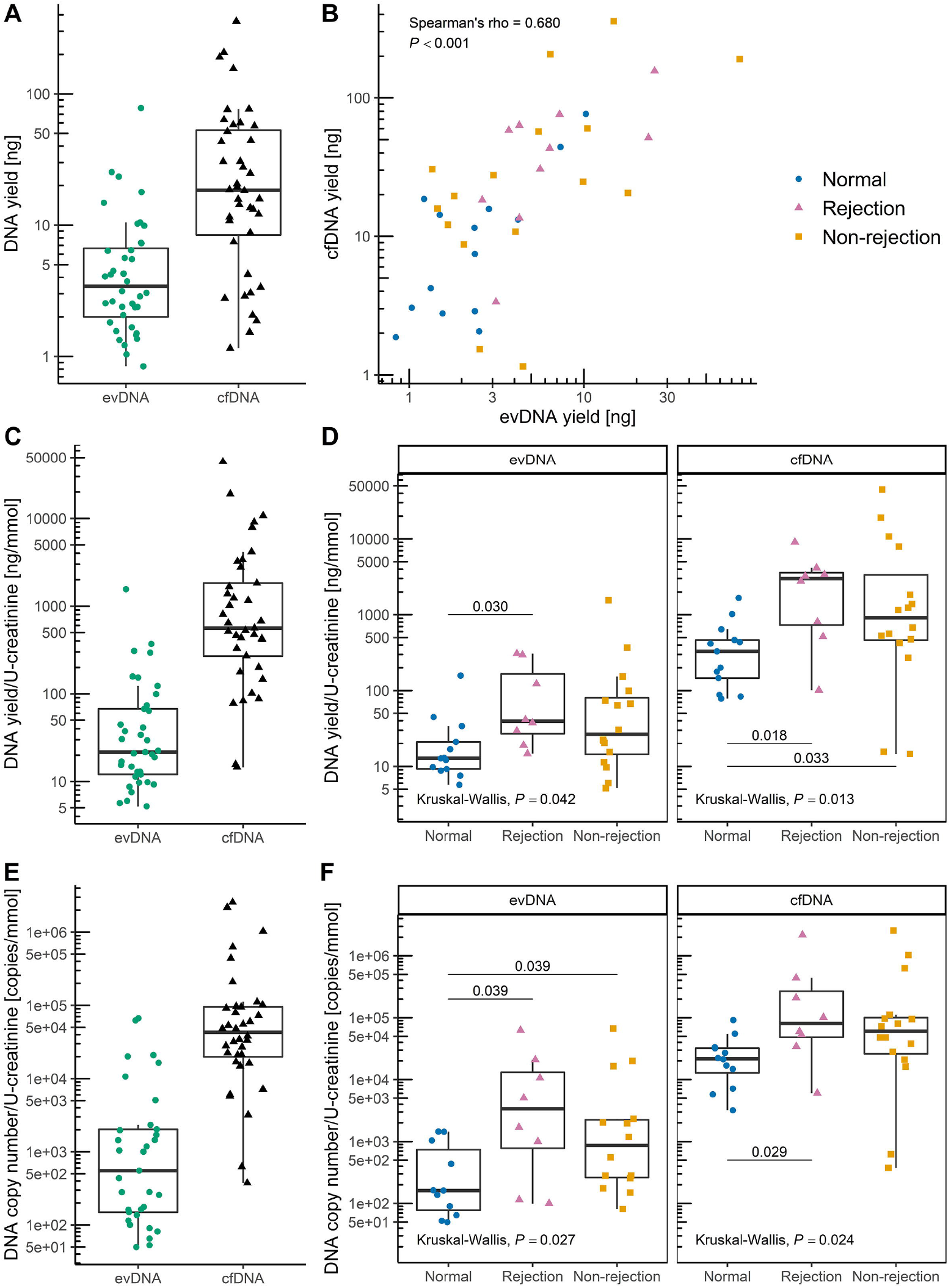
uEV samples of kidney allograft recipients contain DNA. DNA was extracted from uEVs (extracellular vesicle-bound DNA; evDNA) or directly from urine (cell-free DNA; cfDNA) and analyzed for yield by fluorometry and for DNA copy number by detection of RPP30 copies by ddPCR. (A) Distribution of evDNA and cfDNA extraction yields (N = 40) and (B) their correlation (tested with Spearman correlation coefficient). Distribution of DNA yield [ng/mL urine] normalized to U-creatinine [mmol/mL] in (C) the patient cohort (N = 37) and (D) in patient groups with normal histology (N = 13), rejection injury (N = 8), and with non-rejection injury (N = 16). Distribution of DNA copy number normalized to U-creatinine in (E) the patient cohort (N = 33 for evDNA, N = 36 for cfDNA, respectively) and (F) in patient groups with normal histology (N = 11 for evDNA, N = 12 for cfDNA, respectively), rejection injury (N = 8 for evDNA, N = 8 for cfDNA, respectively), and with non-rejection injury (N = 14 for evDNA, N = 16 for cfDNA, respectively). Data of patients with normal histology are presented as blue circles, rejection injury as purple triangles and non-rejection injury as orange squares. Kruskal-Wallis test was used to compare the characteristics between all three patient groups and Mann-Whitney test with Benjamini-Hochberg adjustment for pairwise comparisons between the different groups (*P*-values labeled above the lines connecting different groups in (D, F)).

For further analysis of evDNA and cfDNA, the entire volumes of extracted DNA were first preamplified to obtain enough sample for analysis with multiple ddPCR assays and replicates. In a series of additional experiments, described in more detail in Supplementary Material, we showed that targeted multiplex PCR preamplification maintained the same allele fractions without introducing significant bias (Suppl. Figure 3). We further showed that the preamplification reaction was not inhibited by potential impurities in the DNA extraction eluates, when those were diluted two-times (Suppl. Figure 4). Therefore, we could confidently conclude that preamplified DNA reflects the original quantities of measured DNA fragments and thus the PCR preamplification can be included in the DNA quantification protocol (Suppl. Figure 5). The preamplified evDNA and cfDNA for all samples were next analyzed by ddPCR.

DNA copy number of evDNA and cfDNA samples was determined by measuring the RPP30 copy number (Figure 3E). In four evDNA and one cfDNA samples, RPP30 copy number was too low for reliable quantification, so they were excluded from further analysis. evDNA copy numbers correlated with evDNA yields (ρ = 0.604, *P* < 0.001). evDNA copy number, normalized to U-creatinine, was significantly different between the three patient groups (*P* = 0.027) (Figure 3F). More evDNA copies were measured in patients with rejection injury (*P* = 0.039) and non-rejection injury (*P* = 0.039), than in patients with normal histology. Significance was maintained even when comparing the rejection and non-rejection injury combined with the normal histology group (*P* = 0.007). For cfDNA, the cfDNA copy numbers correlated strongly with cfDNA yields (ρ = 0.918, *P* < 0.001), and also with evDNA copy numbers (ρ = 0.626, *P* < 0.001). cfDNA copy number, normalized to U-creatinine, was significantly different between the three patient groups (*P* = 0.024) (Figure 3F). More cfDNA copies were measured in patients with rejection and non-rejection injury combined than in patients with normal histology (*P* = 0.008).

### 3.4 EV-bound DNA is less fragmented than cfDNA in urine of kidney allograft recipients

The relative degree of fragmentation of evDNA and cfDNA samples was determined by calculating the DNA integrity index, defined as the ratio of RPPH1 (135 bp) to RPP30 (62 bp) amplicon copy numbers. In intact human DNA, the copy number of the two amplicons is the same. However, the more the DNA is degraded, fewer amplicon copies are measured, especially for the longer amplicons. The ratio of long to short amplicons is therefore a good estimate of the DNA fragmentation level (Mouliere et al., 2011). The RPPH1 copy number could not be reliably determined for three evDNA samples, so the evDNA integrity index was not calculated for seven samples (including four in which the RPP30 copy number was already too low). The determined integrity indexes ranged from 0.16 to 0.70 for evDNA, and from 0.13 to 0.44 for cfDNA (Figure 4), but there were no significant differences between the patient groups (*P* = 0.608 for evDNA and *P* = 0.065 for cfDNA). Nevertheless, the cfDNA integrity index was significantly higher in patients with rejection and non-rejection injury combined than in patients with normal histology (*P* = 0.023). Interestingly, cfDNA integrity indexes did not correlate with evDNA integrity indexes (ρ = 0.231, *P* = 0.203), but were significantly lower compared to evDNA in the whole cohort (*P* < 0.001, Figure 4A) as well as in all individual patient groups (Figure 4B) suggesting that cfDNA is more fragmented than evDNA.

**Figure 4:**
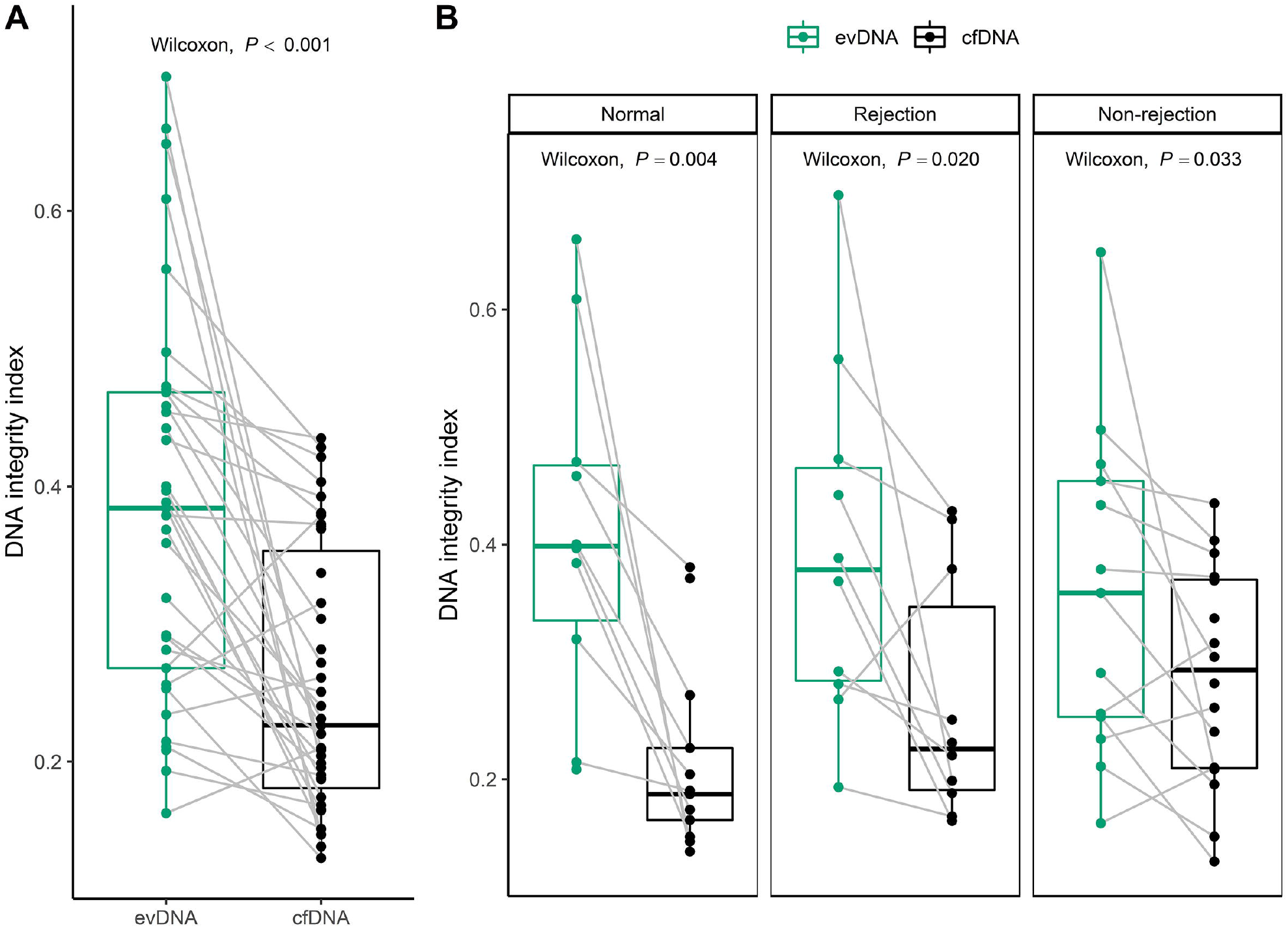
Extracellular vesicle-bound DNA (evDNA) is less fragmented than cell-free DNA (cfDNA) in urine of kidney allograft recipients. DNA integrity index is defined as the RPPH1/RPP30 (long/short amplicon) ratio, as determined by ddPCR. Distribution of DNA integrity indexes of evDNA (green) and cfDNA (black) in (A) the patient cohort (N = 33 for evDNA, N = 39 for cfDNA, respectively) and in (B) patient groups with normal histology (N = 10 for evDNA, N = 13 for cfDNA, respectively), rejection injury (N = 10 for evDNA, N = 10 for cfDNA, respectively), or with non-rejection injury (N = 13 for evDNA, N = 16 for cfDNA, respectively). Gray lines connect paired evDNA and cfDNA samples. The Wilcoxon signed rank test was used for comparison of DNA integrity indexes of related evDNA and cfDNA.

### 3.5 Normalized donor-derived evDNA reflects the kidney allograft rejection phenotype

For analysis of ddDNA content, recipient and donor genomic DNA were genotyped at six SNP loci by real-time PCR and, in ambiguous cases, additionally by ddPCR. For all patients, the recipient and donor genotypes differed in at least two SNP loci, allowing measurement of ddDNA content. Nevertheless, in seven evDNA samples, the number of SNP allelic copies of the donor was too low to allow ddDNA quantification. The ddDNA fraction of evDNA and cfDNA was highly variable, ranging from 2.7% to 99.7% for evDNA, and from 1.8% to 98.6% for cfDNA (Figure 5A). Interestingly, the ddDNA fractions of evDNA and cfDNA correlated strongly (ρ = 0.907, *P* < 0.001), with no significant differences between the patient groups (*P* = 0.303 for evDNA and *P* = 0.113 for cfDNA) (Figure 5B). However, dd-cfDNA fractions were significantly higher in patients with rejection and non-rejection injury combined than in patients with normal histology (*P* = 0.039). When comparing ddDNA copies normalized to U-creatinine, slightly higher ddDNA content was observed in patients with rejection injury, although the differences between patient groups were not statistically significant (*P* = 0.123 for evDNA and *P* = 0.218 for cfDNA) (Figure 5C). When subgroups of rejection cases were compared, significantly more normalized dd-evDNA copies were detected in ABMR than in TCMR (*P* = 0.036) (Figure 5C, Suppl. Table 4).

**Figure 5:**
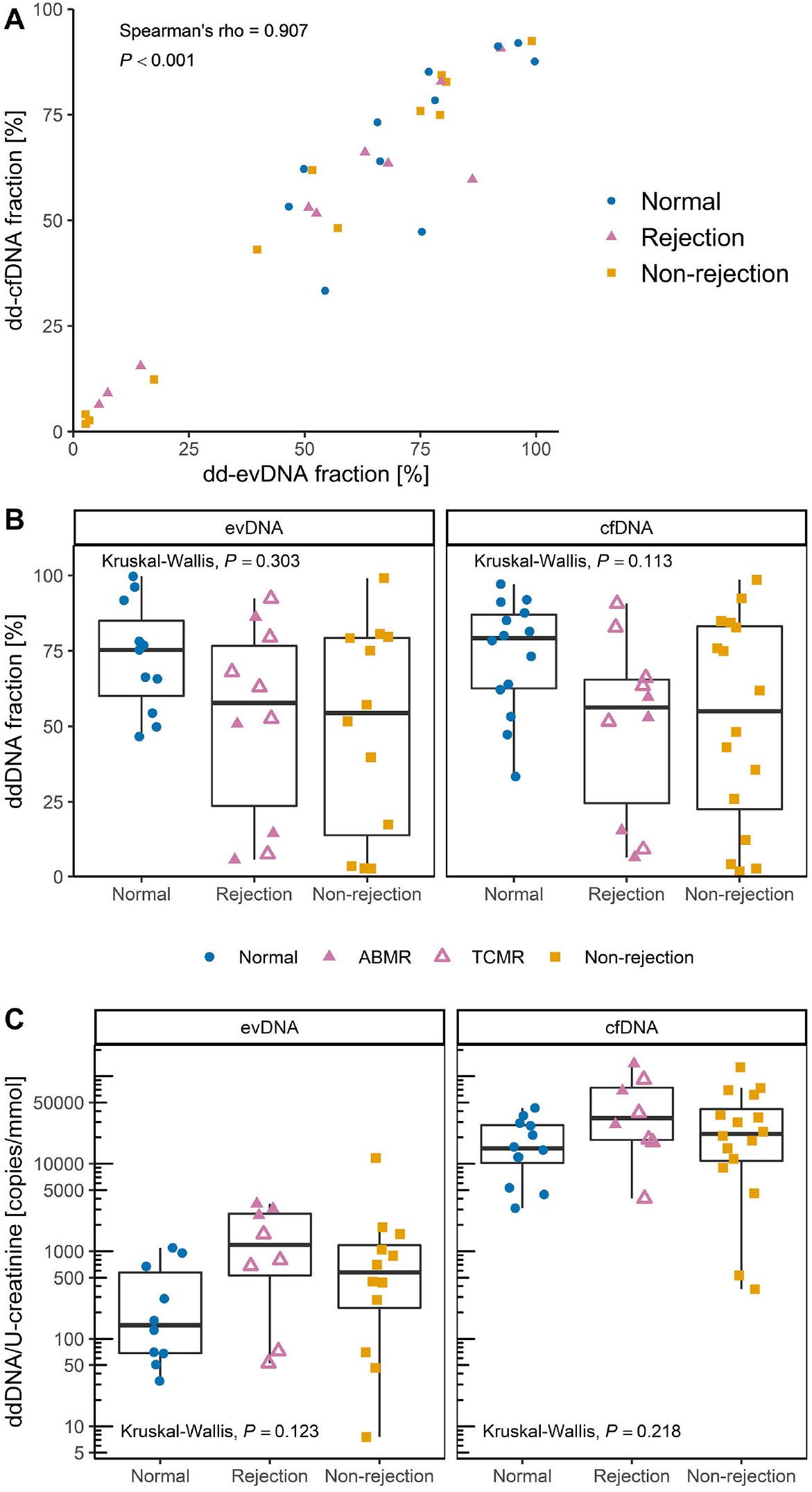
Donor-derived DNA (ddDNA) is detectable in uEV samples of kidney allograft recipients. ddDNA fraction of extracellular vesicle-bound DNA (evDNA) and cell-free DNA (cfDNA) was calculated from ddPCR quantification of at least two single nucleotide polymorphisms (SNPs) that differed between the kidney allograft donor and recipient. The ddDNA copy number was calculated by multiplying ddDNA fraction with DNA copy number. (A) Fractions of dd-evDNA and dd-cfDNA correlate strongly (N = 33; tested with Spearman correlation coefficient). (B) Distribution of ddDNA fractions determined for evDNA (left) and cfDNA (right) in patient groups with normal histology (N = 11 for evDNA, N = 14 for cfDNA, respectively), rejection injury (N = 10 for evDNA, N = 10 for cfDNA, respectively), or with non-rejection injury (N = 12 for evDNA, N = 16 for cfDNA, respectively). (C) distribution of dd-evDNA (left) and dd-cfDNA (right) absolute copy numbers normalized to U-creatinine [mmol/mL] in patient groups with normal histology (N = 10 for evDNA, N = 12 for cfDNA, respectively), rejection injury (N = 8 for evDNA, N = 8 for cfDNA, respectively), or with non-rejection injury (N = 12 for evDNA, N = 16 for cfDNA, respectively). Data of patients with normal histology are presented as blue circles, rejection injury as purple triangles and non-rejection injury as orange squares. In the rejection group, full triangles represent antibody mediated rejection (ABMR), while empty triangles represent T-cell mediated rejection (TCMR). Patients with diagnosis of mixed ABMR and TCMR were grouped within the ABMR subgroup. Kruskal-Wallis test was used to compare the characteristics between all three patient groups.

In summary, DNA co-isolates with urinary EVs and correlates in DNA yield, DNA copy number, and ddDNA fraction with urinary cfDNA, but is less fragmented. More evDNA and cfDNA, as measured by DNA yield, DNA copy numbers or ddDNA fraction (for cfDNA), were recovered from urine samples from patients with rejection and non-rejection injury than from patients with normal histology. Normalized dd-evDNA reflects the kidney allograft rejection phenotype.

### 3.6 DNA is bound to the surface of EVs enriched from urine

The above data indicate that urinary evDNA is better protected from degradation than cfDNA. To test whether evDNA is packaged inside or bound to the surface of EVs, we first treated uEVs of nine study patients (three from each patient group) with DNase I. After treatment, the reaction mixtures were heat inactivated, before proceeding with evDNA extraction, preamplification and characterization. A large reduction in evDNA copy number was observed in DNase-treated compared to untreated uEV samples from the same patient (Figure 6A), as only 2–38% evDNA copies (mean 12%) remained undigested in the analyzed patients’ uEV samples (Figure 6B). To understand whether heat-inactivated DNase retains its activity and contributes in part to the observed reduction in evDNA copies, soybean DNA was added to EV samples in a separate series of experiments and DNase I activity tested at different conditions. 85% of soybean DNA was degraded by DNase during incubation at 75 °C, whereas 59% was degraded when incubated at 37 °C with heat-inactivated DNase (data not shown). These data suggest, that DNase I retains most of its activity even after heat treatment, and could contribute to the observed evDNA degradation. To further evaluate this, we performed additional experiments on three uEV samples, in which DNase was inactivated by addition of denaturing buffer immediately after DNase treatment at 37 °C. Similar to heat inactivation, on average only 14% of the evDNA copies remained undigested after DNase treatment of the uEV samples (Figure 6B). We also confirmed that DNase I did not retain activity after inactivation by denaturing buffer (data not shown).

**Figure 6:**
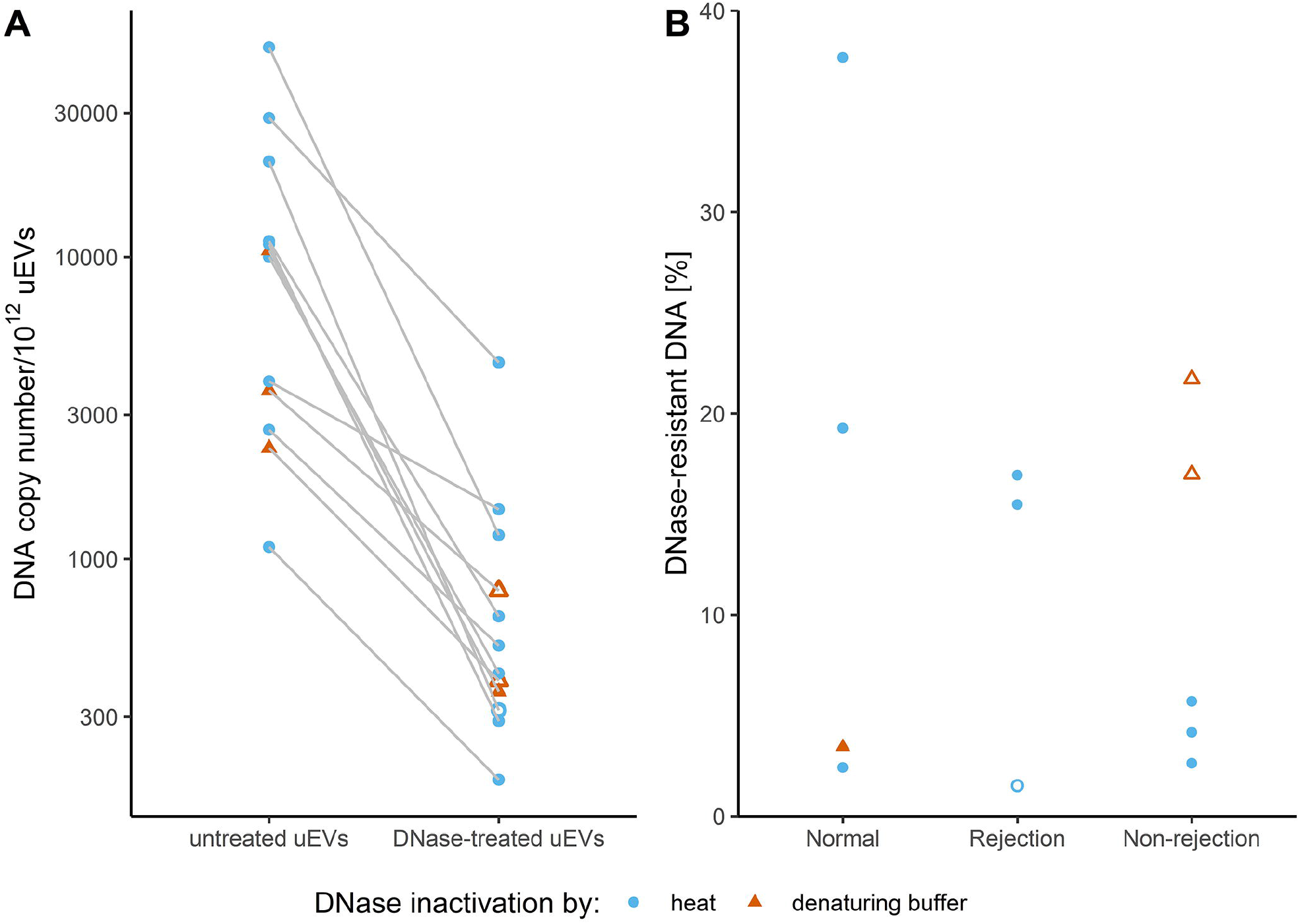
DNase treatment degrades most of the DNA in uEV samples of kidney allograft recipients. DNA copy number in uEV samples, isolated from selected patients with normal histology, rejection injury or non-rejection injury, without or with DNAse treatment was determined by measuring the RPP30 copies by ddPCR. DNase was inactivated by heat (light blue circles) or by immediate DNA extraction (red triangles). Empty circles and triangles represent less reliable measurements of DNA copies in EV samples, as concentration of DNA after DNase treatment was very low. (A) DNA copy numbers normalized to 10^12^ uEVs in untreated and DNase-treated uEV samples. (B) DNase-resistant fraction of DNA in treated uEV samples.

This suggests that most of evDNA is sensitive to degradation by DNase I, but as shown here, evDNA is also less fragmented compared to cfDNA in urine. To test whether evDNA is protected from degradation in urine by binding to the surface of EVs, we labelled DNA in representative uEV samples from patients with normal histology, rejection injury, or non-rejection injury and analyzed them using TEM (Figure 7). We used antibodies against dsDNA, which in turn were recognized by IgG antibodies conjugated to gold particles. Representative micrographs show that at least some of the uEVs in analyzed samples were labelled with aggregated gold particles, indicating binding of dsDNA to the surface of EVs (Figure 7B). On the other hand, gold particles were not present in representative uEV samples where incubation with antibodies against dsDNA was omitted (Figure 7A). Interestingly, despite the absence of dsDNA-specific antibodies, the uromodulin filament was labelled with gold particles in one of the samples (Suppl. Figure 6), as previously noted (Rhodes et al., 1993). In summary, DNA is bound to the surface of EVs enriched from urine, independent of the patients’ allograft injury status.

**Figure 7.**
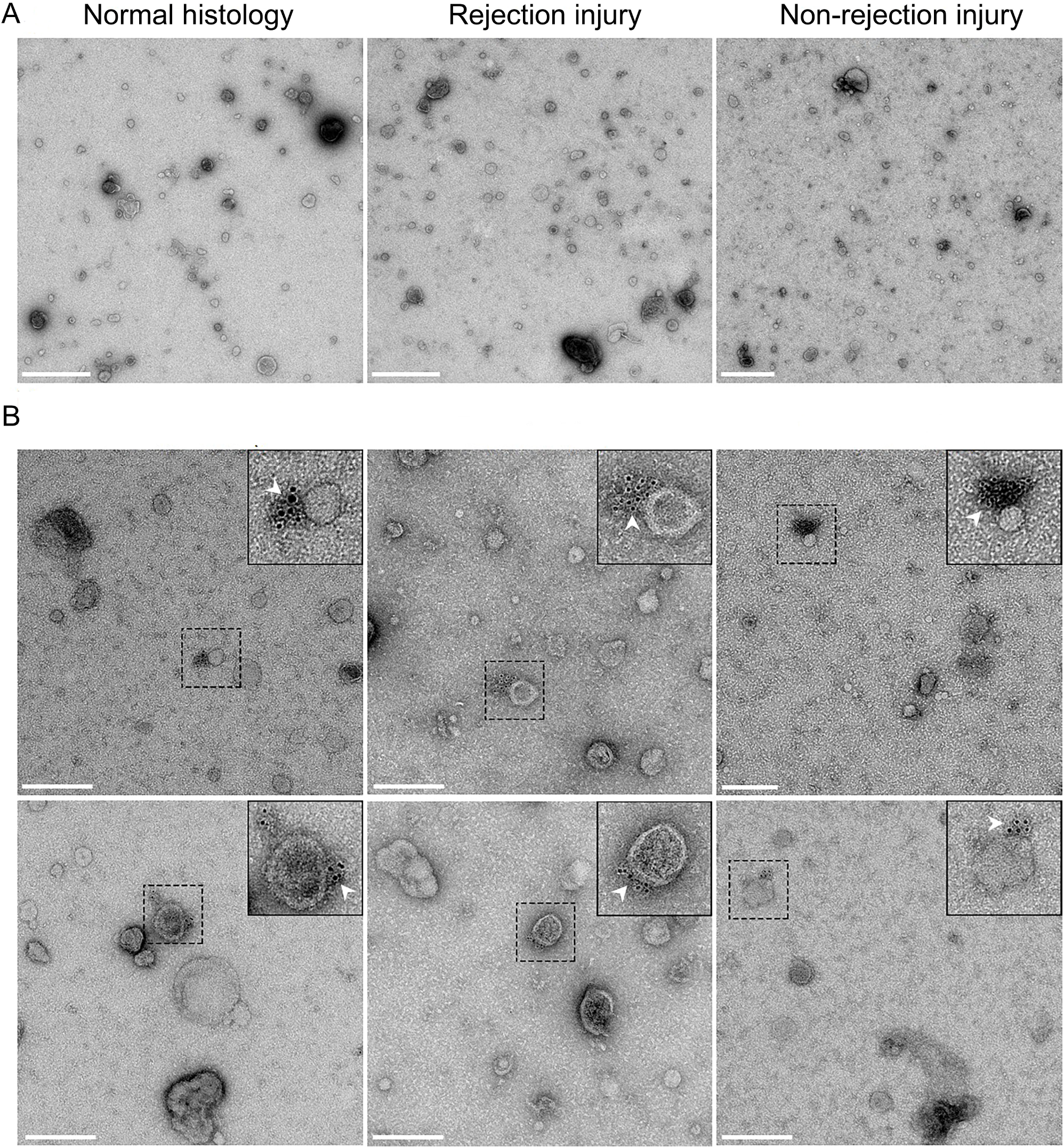
DNA in uEV samples of kidney allograft recipients is bound to the surface of extracellular vesicles. (A) Transmission electron micrographs of representative negative-stained uEV samples from the normal histology (left), rejection injury (middle) and non-rejection injury (right) patient groups. Scale bar: 500 nm. (B) Micrographs of two representative fields of vision of uEV samples of the three patients groups, labelled with antibodies against dsDNA and IgG conjugated with gold particles. White arrows mark colloidal gold particles (6 nm), indicating the location of the DNA. Scale bar: 200 nm.

### 3.7 evDNA characteristics associate with clinical parameters of kidney allograft recipients

Finally, to evaluate the potential of urinary evDNA as a biomarker of kidney allograft injury, the association of evDNA characteristics with patients’ clinical characteristics was assessed. Associations with categorical variables are presented in Table 2 and Suppl. Table 4, and with continuous variables in Suppl. Table 5. Among the clinical characteristics assessed, normalized evDNA copy numbers were significantly increased (*P* = 0.025) in patients with for-cause biopsy compared to those with surveillance biopsy. Normalized evDNA yield, evDNA copy number, and dd-evDNA copy number were significantly higher in patients with an interstitial inflammation score > 0, compared with those with a score 0 (all *P* < 0.050, Table 2). Banff score 0 represents the absence or evidence of less than 10% inflammation in the specific region of the biopsy specimen, whereas scores > 0 indicate inflammatory lesions graded with scores 1, 2, or 3. As expected, higher interstitial inflammation scores were detected in patients with allograft rejection injury, particularly in those with TCMR (Figure 8A). For the combined parameters of glomerulitis and peritubular capillaritis, normalized evDNA yield, evDNA copy number and dd-evDNA copy number were also significantly higher in patients with Banff lesion scores > 0 compared with those with score 0 (all *P* < 0.050). As expected, higher combined scores for glomerulitis and peritubular capillaritis were observed in patients with ABMR (Figure 8B). A similar trend was observed for inflammation in areas of fibrosis, where normalized evDNA copy numbers were higher in patients with Banff lesion scores > 0 (*P* = 0.040). Higher inflammation in areas of fibrosis score was observed in patients with rejection or non-rejection injury, compared to patients with normal histology (Figure 8C). Interestingly, several evDNA characteristics differed with respect to patient sex. The dd-evDNA fraction was lower (*P* < 0.001), whereas the evDNA integrity index (*P* = 0.025) and normalized evDNA yield (*P* = 0.026) were higher in females than in males (Table 2). Similarly, uEVs were larger in females than in males (Suppl. Table 3; mean size (*P* = 0.004), mode size (*P* = 0.043), median size (*P* = 0.009)).

**Table 2:**
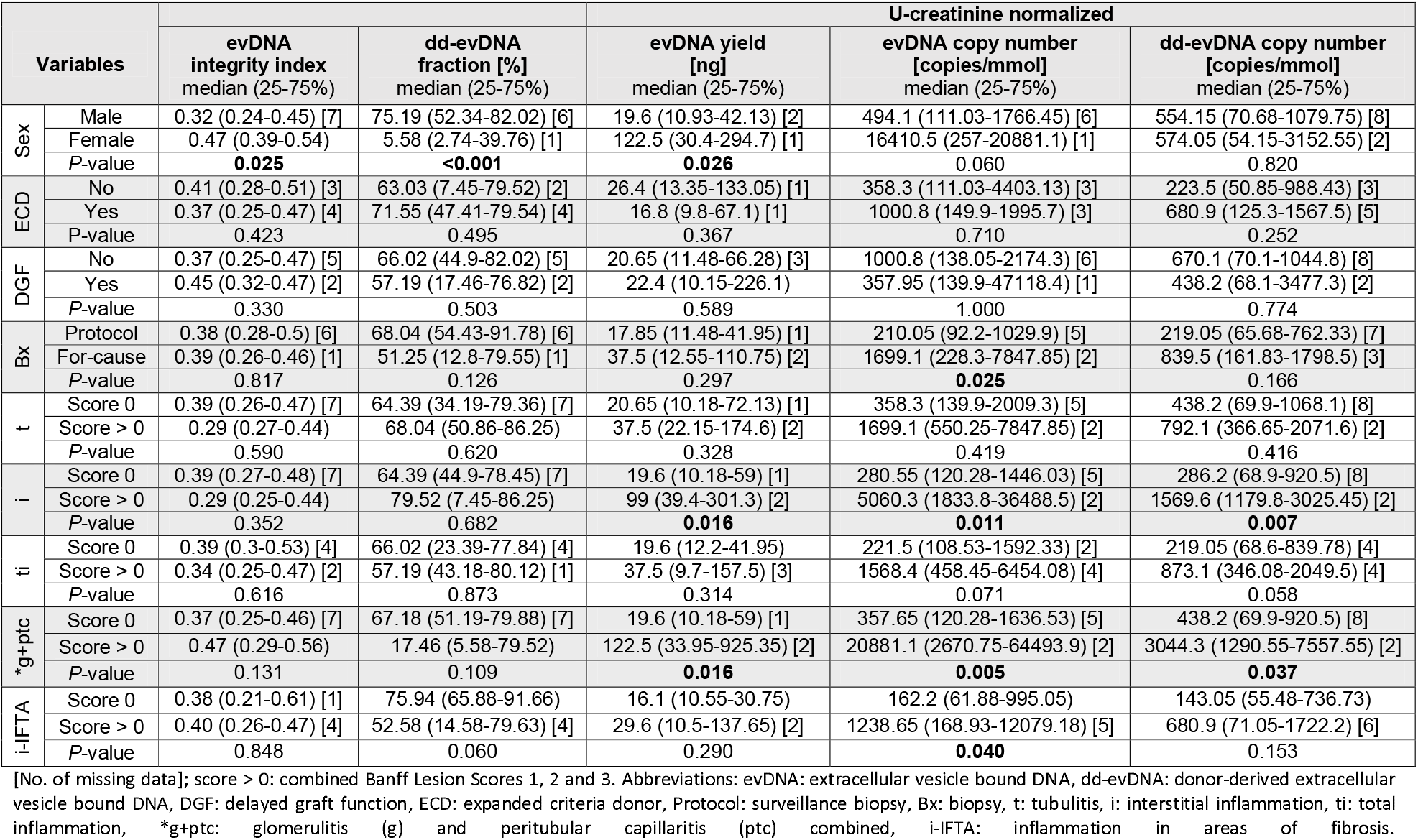
Association of evDNA characteristics with kidney allograft recipients’ clinical and histological parameters (categorical variables).

**Table 3:**
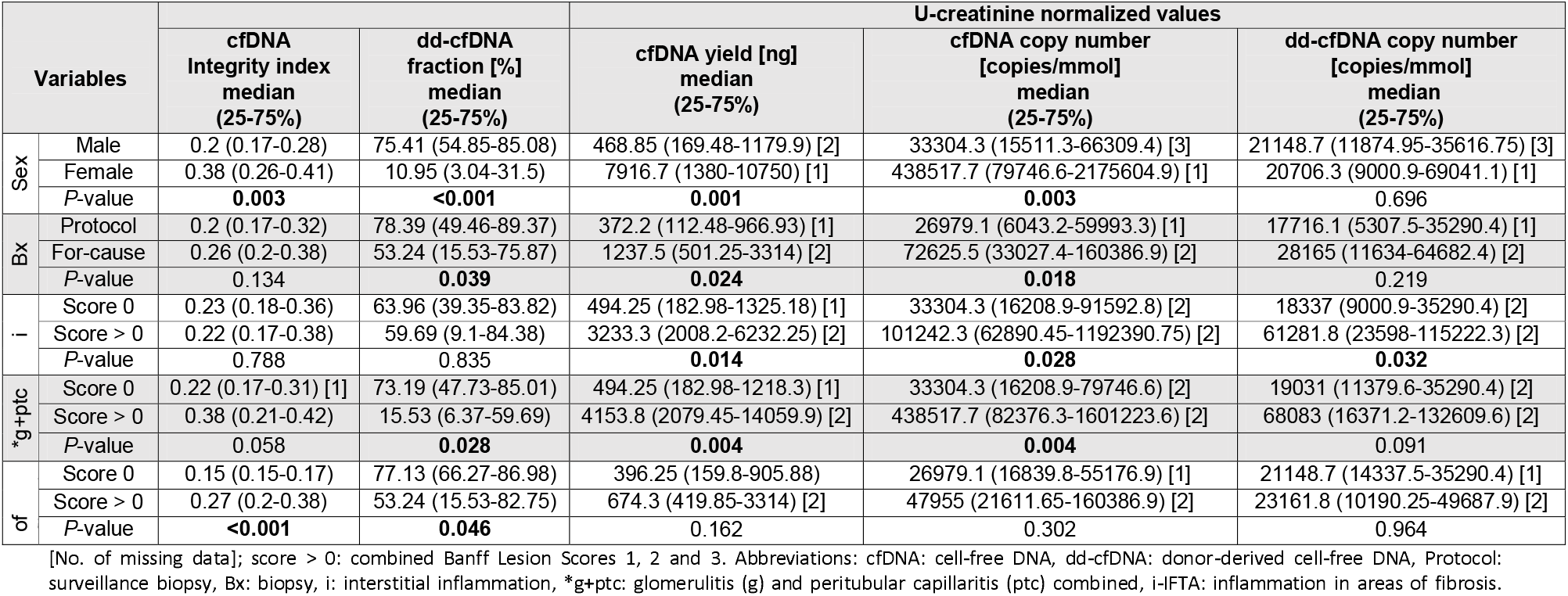
Association of cfDNA characteristics with kidney allograft recipients’ clinical and histological parameters (categorical variables). Only cfDNA parameters that are significantly associated with clinical variables are presented.

**Figure 8:**
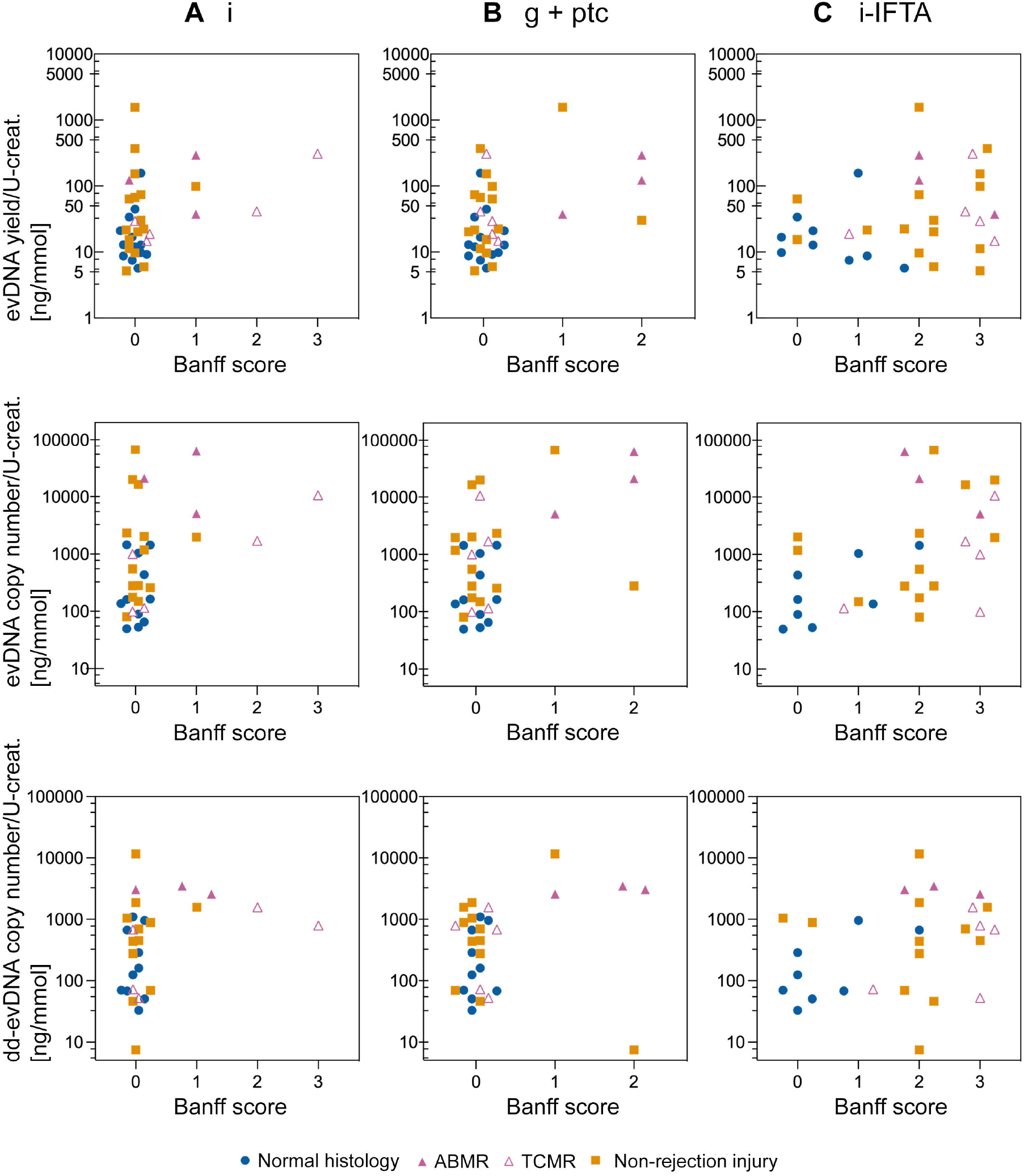
Distribution of normalized evDNA yield, evDNA copy number and dd-evDNA copy number for each value of the selected Banff parameters. Each data point represents individual patient, with blue circles, purple triangles and orange squares denoting data of patients with normal histology, rejection injury (antibody mediated rejection; ABMR - full, T-cell mediated rejection; TCMR - empty) or non-rejection injury, respectfully. Banff score 0 is indicative of absence or detection of less than 10% inflammation in the specific region of the biopsy sample, while inflammatory lesions are graded with scores 1, 2 or 3 depending on the severity. Abbreviations; i: interstitial inflammation, g+ptc: patients with glomerulitis (g) and peritubular capillaritis (ptc) combined, i-IFTA: inflammation in areas of fibrosis, evDNA: extracellular vesicle bound DNA, dd-evDNA: donor-derived extracellular vesicle bound DNA.

To evaluate the relevance of evDNA as a biomarker of kidney allograft injury in comparison to cfDNA from urine, association of cfDNA characteristics with patients’ clinical characteristics was performed. (Table 3, Suppl. Table 6). Similar to evDNA, urine cfDNA measured characteristics were significantly associated with sex, the type of biopsy, and the level of interstitial inflammation, combined glomerulitis and peritubular capillaritis, and inflammation in areas of fibrosis (Table 3). Despite observed similarities, additional cfDNA (or evDNA) characteristics correlated with certain patients’ clinical characteristics. In patients with for-cause biopsy, dd-cfDNA fraction and normalized cfDNA yield significantly differed compared to patients with surveillance biopsy (*P* = 0.039 and *P* = 0.024 respectively). For the combined parameters of glomerulitis and peritubular capillaritis, dd-cfDNA fraction was significantly elevated in patients without inflammatory lesions (*P* = 0.028), while only a trend was observed for the normalized dd-cfDNA copy number (*P* = 0.091), as opposed to evDNA. Inflammation in areas of fibrosis showed to be most variable, as cfDNA integrity index and dd-cfDNA fraction significantly differed in the presence or absence of inflammatory lesions (*P* < 0.001 and *P* = 0.046, respectively), while normalized cfDNA copy number did not associate with lesions (*P* = 0.302), as opposed to evDNA. To conclude, urine evDNA has a great potential as biomarker of kidney allograft injury, but can be interchanged with urine cfDNA.

## 4 Discussion

The purpose of this study was to investigate DNA as urinary EV cargo, using kidney transplantation as a model to investigate its biomarker potential. To this end, we enriched EVs from the urine of well-characterized kidney transplant recipients and explored the EV-bound DNA and its association to patients’ clinical characteristics. Using DNase treatment and immunogold labelling TEM, we showed for the first time that DNA is mostly bound to the surface of EVs in urine. Although urinary evDNA and cfDNA correlated in several characteristics, evDNA was less fragmented as measured by the DNA fragmentation index. Urinary EVs from patients with kidney allograft injury were significantly larger and had higher amounts of bound DNA, as measured by normalized evDNA yield and evDNA copy number, compared to patients with normal histology. In addition, urinary evDNA characteristics reflect the kidney allograft injury, since they were significantly associated with the type of biopsy and the extent of interstitial inflammation, combined glomerulitis and peritubular capillaritis, and inflammation in areas of fibrosis. Importantly, normalized dd-evDNA copies differed significantly between the two rejection types, ABMR and TCMR. Thus, evDNA has a great potential as biomarker for kidney allograft injury, and specifically for allograft rejection.

An important finding of our study is the first experimental evidence that dsDNA is bound to the surface of EVs in urine. evDNA was mostly unprotected from DNase I degradation, but localized to the EV surface in immunogold DNA-labelling TEM. This may explain the previous observation that DNA was co-isolated with microvesicles from human and rat urine, which was at the time of discovery designated as contaminant (Miranda et al., 2010). Nevertheless, up to 38% of evDNA remained undigested in our study, suggesting that evDNA might also be incorporated inside EVs to small extent or protected from degradation as part of exomeres (Jeppesen et al., 2019), so far not yet detected in urine (Zhang et al., 2018). Studies on EVs from blood plasma or serum (Kahlert et al., 2014; Lázaro-Ibáñez et al., 2014; Vagner et al., 2018) and cell cultures (Lázaro-Ibáñez et al., 2019; Németh et al., 2017) similarly showed that DNA was mostly bound to the surface of small EVs, whereas it was incorporated inside larger EVs. In those studies, DNA EV-localization was largely determined by DNase degradation assay. One study demonstrated colocalization of DNA dye with EV-typical proteins CD63 and CD81 using super-resolution imaging of single EVs (Maire et al., 2021), whereas another study demonstrated localization of DNA dye with EVs labelled with annexin V, anti-CD63 using flow cytometry (Németh et al., 2017). Moreover, our observations on evDNA localization in the context of kidney transplantation are valuable, because most of what is known about evDNA has so far been studied in the context of cancer or cancer cell lines, with characteristic genome instability not seen in most other pathologies (Lázaro-Ibáñez et al., 2019; Lee et al., 2018; Vagner et al., 2018).

The mechanism of DNA binding to the uEV surface, i. e., is DNA bound to EV surface during endosomal biogenesis (Jeppesen et al., 2019) or is it a consequence of DNA binding to cell surface in the process of plasma membrane invagination (Tamkovich & Laktionov, 2019), is not known. But as shown here, urinary evDNA and cfDNA correlated in DNA yield, DNA copy number, and ddDNA fraction, implying that DNA may have the same source in both cases. At the molecular level, studies on DNA binding to the cellular membrane surface show that DNA-membrane binding can be promoted through electrostatic interactions (Laktionov et al., 2004), direct binding of DNA by specific membrane receptors (Rykova et al., 2012), or indirect binding through histones (Kubota et al., 1990). As EVs share characteristics with parental cells, similar mechanism could exist. This is supported by the following observations: (i) EV adhesive capabilities are influenced by their structure (Kawamura et al., 2017), (ii) the cell-surface DNA-binding receptors MAC-1, Ku80, and Ku70 have been identified in uEV proteomic studies (Silvers et al., 2017; Wang et al., 2012), (iii) histones have been detected on EVs (Németh et al., 2017) or DNA-bound to EVs (Lázaro-Ibáñez et al., 2019; Takahashi et al., 2017). Regardless, based on observation that DNA weakly linked to cellular surface can be easily detached by 5 mM EDTA (Laktionov et al., 2004), DNA association to the uEVs was stronger in our study, because 8 mM final concentration of EDTA was added to the urine during the processing step to prevent polymerization of uromodulin (Kobayashi & Fukuoka, 2001). Future in-depth characterization of evDNA and cfDNA sequences, epigenetic modifications and DNA-bound proteins may help answer these questions.

Importantly, urinary EVs in our cohort reflect kidney allograft rejection and non-rejection injury. The uEVs were significantly larger in the rejection and non-rejection injury groups compared with the normal histology group. Normalized DNA yields and evDNA copy number in the three groups were statistically significantly different, with more urinary evDNA obtained in patients with rejection injury and non-rejection injury when compared to normal histology. Higher normalized evDNA copy numbers also significantly correlated with for-cause biopsy, performed in patients with increased serum creatinine or proteinuria. In addition, higher normalized evDNA yield, evDNA copy number, and dd-evDNA copy number correlated with inflammation in the kidney allograft (specific Banff scores > 0). Previous studies have shown that subclinical inflammation is a relatively common finding in surveillance allograft biopsies, identifying a high-risk group of kidney recipients for poor subsequent outcomes (Seifert et al., 2021). Other studies focusing on kidney injury observed larger uEV size in properly controlled diabetic patients compared with healthy controls (Freeman et al., 2018), whereas no changes in uEV size were observed when comparing different groups of patients with acute or chronic glomerular disease (Dimuccio et al., 2020). The uEV proteins neutrophil gelatinase-associated lipocalin (Alvarez et al., 2013), CD133 (Dimuccio et al., 2014), CD3 (Park et al., 2017), and aquaporin-1 (Sonoda et al., 2009), as well as a specific set of uEV RNAs, have been previously associated with kidney injury (El Fekih et al., 2021), but our study is, to our knowledge, the first to correlate urine evDNA with kidney injury.

Monitoring kidney function using urinary evDNA is particularly useful in allogenic kidney transplants because their genomic sequence differs from that of the transplanted patient. Thus, we can use the differences in genotype to specifically track the DNA released from the allograft (ddDNA) due to apoptosis, necrosis, or active release (Oellerich et al., 2021; Sigdel et al., 2013; Zhang et al., 1999). Here, we have successfully measured dd-evDNA fraction and dd-evDNA copy number in the urine of our kidney transplant patient cohort, using our recently developed ddPCR method for determining dd-cfDNA in plasma (Jerič Kokelj et al., 2021). Because a recent study reported a significant effect of variable water-load on the concentration of uEVs (Blijdorp et al., 2021), we adjusted the uEV and DNA values to urine creatinine to account for interpatient variability in urine concentration (Erdbrügger et al., 2021). The ddDNA fraction of evDNA was highly variable, ranging from 2.7% to 99.7%, but no significant differences were detected among the three patient groups. When absolute ddDNA levels were evaluated, a trend toward more normalized dd-evDNA copies was observed in patients with rejection injury. Importantly, significantly more normalized dd-evDNA copies were detected in ABMR than in TCMR. Overall, it appears that absolute quantification of dd-evDNA in urine is more informative for allograft rejection compared to the dd-evDNA fraction, similar to the study by Oellerich et al. (2019). This could be due to fluctuations in recipient DNA levels due to infection, exercise, or psychological stress, which can damage the recipient’s own cells, resulting in increased levels of recipient extracellular DNA that affect dd-cfDNA fraction calculations (Oellerich et al., 2021). Based on the above, urinary dd-evDNA has great potential as a biomarker for kidney allograft rejection, particularly in differentiating between ABMR and TCMR. Discriminating rejection from no rejection status and ABMR from TCMR phenotype can help to refine the diagnosis and provide the clinicians with useful information for clinical management decisions.

Several large, multicenter cohort studies support the use of plasma dd-cfDNA levels to monitor kidney allograft rejection, performing most robustly for discriminating ABMR (Bloom et al., 2017; Gielis et al., 2020; Halloran et al., 2022; Huang et al., 2019; Oellerich et al., 2019; Sigdel et al., 2018; Whitlam et al., 2019). To date, three commercial blood-based dd-cfDNA assays have become available for clinical use (Filippone & Farber, 2021; Paul et al., 2021). Our study on urinary cfDNA is one of the few studies investigating the association between urinary dd-cfDNA and kidney injury and rejection. Sigdel et al. had previously shown that urinary dd-cfDNA is significantly higher in patients with acute rejection than in patients with stable kidney allograft (Sigdel et al., 2013), whereas urinary cfDNA analysis is part of the multimarker test for early detection of kidney allograft rejection (Watson et al., 2019; Yang et al., 2020). Here, we showed that the ddDNA fraction of cfDNA varied widely, similar to evDNA, with no significant differences detected among the three patient groups. As with dd-evDNA, normalized ddDNA levels showed a trend towards more dd-cfDNA copies in patients with rejection injury. While dd-cfDNA fractions reliably distinguished patients with allograft injury (rejection or non-rejection) from patients with normal histology, differentiation of two rejection subgroups based on normalized dd-cfDNA copies was not possible.

Although urinary cfDNA and evDNA in our study show similar trends in the detection of kidney allograft injury, evDNA is less fragmented based on the measured DNA integrity index. Higher integrity index of evDNA potentially enables less restrictive ddPCR (or any other PCR) assay design in terms of amplicon length, which is highly and progressively limited when dealing with more degraded DNA. But at the same time, the total DNA quantity needs to be considered as well. Thus, the benefit of evDNA having higher integrity index is in a way balanced with higher cfDNA yield on the other side, as the poor evDNA yield is on the limit for reliable quantification. Nevertheless, unlike cfDNA, EVs carry a variety of biological signals, which allows multiplexing and offers the possibility of identifying the origin of cellular cargo, which in turn could improve detection specificity (Chennakrishnaiah et al., 2019). Furthermore, EVs protect their molecular cargo from degradation, allowing relevant signals to be recovered even from archival samples (Chennakrishnaiah et al., 2019). In biomarker studies of pancreatic ductal adenocarcinoma, a higher rate of KRAS mutations was detected in evDNA than in cfDNA (Allenson et al., 2017), and only evDNA levels correlated with patient outcomes (Bernard et al., 2019). Thus, evDNA and cfDNA are mostly interchangeable biomarkers in our study, but evDNA was shown to outperform cfDNA in cancer, especially when detecting specific mutations or other nucleotide changes.

Interestingly, the analysis showed some significant differences with respect to the sex of the patients. In contrast to reports that urine from female kidney donors is more enriched in EVs (Turco et al., 2016), we did not detect differences in uEV concentration. However, our results show larger DNA fragments and abundant DNA load in the female patients. In males, the dd-evDNA and dd-cfDNA fractions were significantly increased, whereas the total DNA copy number was higher in females than in males. Overall, this implies the necessity to investigate the influence of sex on EV characteristics.

The advantage of our study is that it is a single-center study in which patients were treated according to the same and well-established treatment protocol. Importantly, clinical samples were collected according to the same protocol and at fixed time points, which may have helped reduce the impact of preanalytical variables on the measured characteristics. In addition, histologic assessment of allograft injury was performed by a dedicated histopathologist, reducing the possibility of interobserver variability in interpretation of results. There are a few limitations to this study. First, the total number of rejection cases included is small because of the effective immunosuppressive therapy and long-term survival of allografts in transplanted patients. Second, we have not collected serial urine samples, which prevented us from testing how early the urinary evDNA and cfDNA can predict allograft injury and rejection before the biopsy. Finally, our study has not included an independent validation cohort.

In conclusion, our results show that most of the DNA bound to EVs in urine is located on the EV surface. Although urinary evDNA and cfDNA correlate in several characteristics, evDNA appears less fragmented and therefore allows for less restricted ddPCR analysis, given that adequate volume of urine is collected to achieve sufficient evDNA extraction yield. In addition, evDNA reliably reflects kidney allograft injury and the severity of inflammation in specific compartments of kidney allograft/biopsy specimens. Overall, evDNA is a promising biomarker for kidney allograft injury, particularly for allograft rejection and differentiation between the two types of rejection, ABMR and TCMR. Further validating studies with prospectively collected samples would help confirm our findings in larger patient cohorts and evaluate the potential of these urinary biomarkers for early detection of kidney allograft injury and rejection.

## Supporting information

MISEV2018 Checklist

REMARK for EQUATOR network Checklist

## Data Availability

All data produced in the present work are contained in the manuscript and supplementary material.

## Geolocation information

46.050980777270254, 14.518433998551663

## Acknowledgements

We thank mag. Tina Demšar for soybean gDNA and Le1 assay for its quantification.

## Supporting material

additional file appended (Suppl. Figure 1-6, Suppl. Table 1-6 and complementary description of results).

