## Supplemental data for "Extracellular vesicle-bound DNA in urine is indicative of kidney allograft injury"

**Supplementary material:**

**S1 Size profiles of enriched uEVs for each patient group, as determined by NTA analysis**

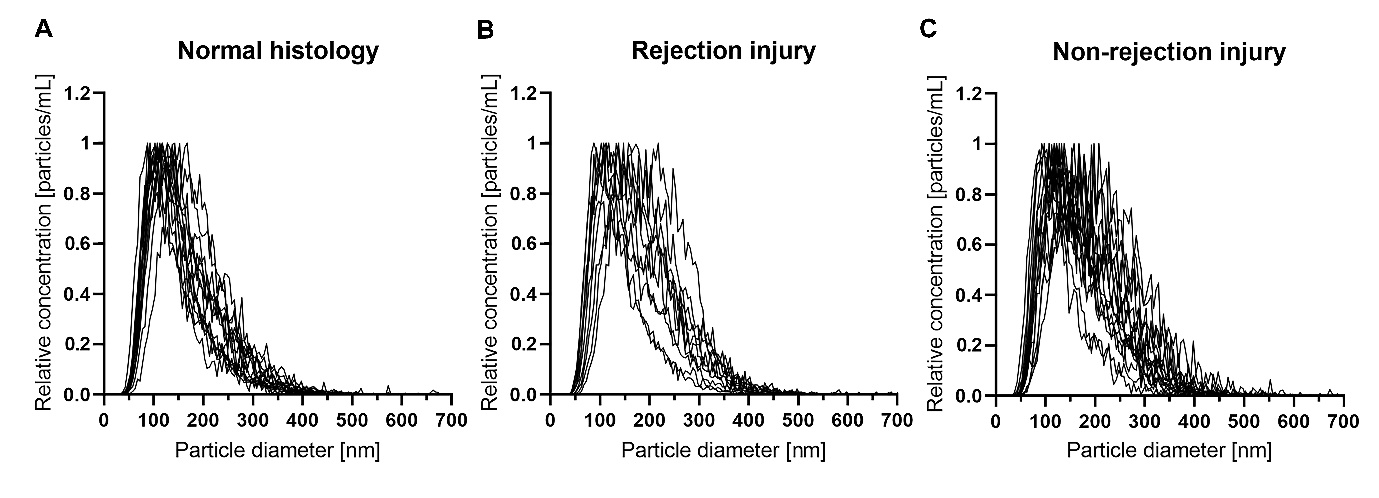

**Supplementary Figure 1: Size profiles of enriched uEVs as determined by NTA analysis.** Size profiles for each included patient with (A) normal histology, (B) rejection injury or (C) non-rejection injury are presented. Y-axis presents uEV concentrations normalized to maximal uEV concentration value in each sample.

**S2 Comparison of five commercially available kits for cfDNA extraction from urine**

cfDNA was extracted from 4 mL of the same pooled urine sample with five commercially available kits, and DNA concentration determined fluorometrically. The key characteristics of the tested kits are presented in Suppl. Table 1. Manual extraction with Quick-DNA Urine Kit resulted in high yield and also cost efficiency and the protocol was not too laborious. The kit’s biggest drawback is unavailability of automated procedure. Automated protocols for other kits were tested using the KingFisher instrument for automated purification (Thermo Scientific), but resulted in lower cfDNA yields than manual protocols. Nevertheless, optimization of the automated protocols might in the future improve cfDNA extraction efficiency. cfDNA, isolated by NextPrep-Mag Urine cfDNA Isolation kit, inhibited ddPCR reaction and resulted in lower fluorescence amplitude of positive droplets.

**Supplementary Table 1**: Key characteristics of the five tested commercially available kits for cfDNA extraction from urine

| **Characteristics** | **Commercial kits for cfDNA isolation** | | | | |
| --- | --- | --- | --- | --- | --- |
|  | Apostle | MagMAX | NextPrep-Mag | Quick-DNA | QIAamp |
| cfDNA yield | +++ | ++ | + | +++ | + |
| Handling | medium | demanding | easy | medium | Easy |
| Automatization | + | + | + | - | + |
| Price per sample | €€ | €€€ | € | € | €€ |
| Max volume of urine | 5 mL | 10 mL | 20 mL | 40 mL | 4 mL |
| Other remarks | Magnetic stands required | Magnetic stands required | Magnetic stands required, not recommended for ddPCR | Fume hood required | Vacuum pump required; cleaning and decontamination of the pump required |

Abbreviations; Apostle, Apostle MiniMax High Efficiency cfDNA Isolation Kit (Beckman Coulter); MagMAX, MagMAX Cell-Free DNA Isolation Kit (Applied Biosystems); NextPrep-Mag, NextPrep-Mag Urine cfDNA Isolation Kit (Perkin Elmer); Quick-DNA, Quick-DNA Urine Kit (Zymo Research); QIAamp, QIAamp Circulating Nucleic Acid Kit (Qiagen)

The Apostle and Quick-DNA kits had similar performance in studied cfDNA characteristics (Suppl. Figure 2), while the MagMAX resulted in lower yield and integrity index of urine cfDNA. Based on the size profiles, MagMAX was able to extract shorter cfDNA compared to Apostle and Quick-DNA kits, but was less efficient in extracting the higher molecular weight cfDNA.

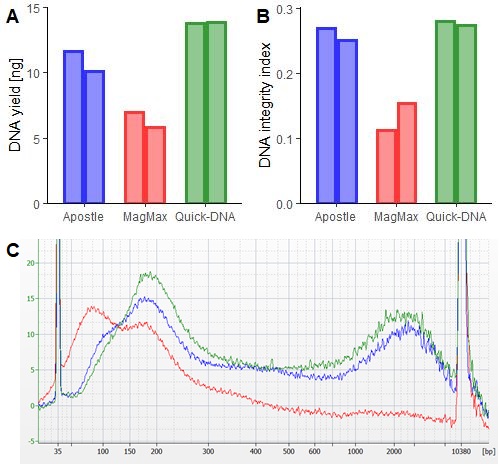

**Supplementary Figure 2**: Comparison of (A) yield, (B) integrity index and (C) size profile of cfDNA extracted from 4 mL of pooled urine with the three best performing commercially available kits: Apostle (blue), MagMAX (red), or Quick-DNA (green). DNA integrity index is defined as the ratio of RPPH1 (135 bp) to RPP30 (62 bp) amplicon copy numbers, as determined by ddPCR. For size profiling, 1 µL of urine cfDNA was analyzed on Bioanalyzer 2100 using DNA High Sensitivity Kit (both from Agilent). Apostle, Apostle MiniMax High Efficiency cfDNA Isolation Kit (Beckman Coulter); MagMAX, MagMAX Cell-Free DNA Isolation Kit (Applied Biosystems); Quick-DNA, Quick-DNA Urine Kit (Zymo Research).

**S3 Validation of DNA preamplification reaction**

Before analyzing clinical samples, we tested reliability of the targeted multiplex PCR preamplification of six SNP loci. First, we prepared 3 or 5% (w/w) mixtures of two genomic DNAs with genotypes that differed in all six tested SNPs, to mimic low levels of donor derived DNA (spike-in DNA) in the background of recipient’s DNA (background DNA), as observed in patients with kidney transplant. The background DNA genotype was rs1707473 G/G, rs2691527 C/C, rs7687645 T/T, rs1420530 T/T, rs9289628 C/C, rs6070149 T/T, while the spike-in DNA genotype was rs1707473 T/G, rs2691527 T/C, rs7687645 C/T, rs1420530 T/C, rs9289628 T/T, rs6070149 C/C. We added 6.8 ng of DNA mixture into the preamplification reaction, which corresponds to approximately 2000 DNA copies or haploid genome equivalents. Thus, minor allele was present at about 60 or 30 copies per preamplification reaction in the 3%, and 100 or 50 copies per preamplification reaction in the 5% genomic DNA mixture for homozygous or heterozygous state, respectively.

Next, we used ddPCR to determine the allele fractions of all 6 SNPs (rs1707473, rs2691527, rs7687645, rs1420530, rs9289628, rs6070149) in PCR-preamplified genomic DNA mixtures, and compared it to those determined for the same DNA mixtures without preamplification (see Materials and Methods for ddPCR protocol). All ddPCR reactions were performed in triplicates. Importantly, allele fractions of tested SNPs were comparable in genomic DNA mixtures with or without PCR preamplification (Suppl. Figure 3), as no significant change in mean allele fractions was observed after preamplification (*P* = 0.150 for the 3% and *P* = 0.337 for the 5% DNA mixture). As expected, coefficient of variation was larger for the mean allele fraction in the 3% compared to 5% DNA mixture. Calculated efficiency of preamplification for all tested amplicons, which is defined as the ratio between ddPCR determined copy number after preamplification to theoretically expected copy number after preamplification, was 84 ± 17% (mean ± sd).

To support this data, ratio of RPPH1 to RPP30 amplicon copy numbers (defined also as DNA integrity index) was determined by ddPCR in PCR-preamplified genomic DNA mixtures, and compared to the determined ratio in DNA mixtures without preamplification. Importantly, the RPPH1/RPP30 ratio did not change significantly after preamplification (*P* = 0.100).

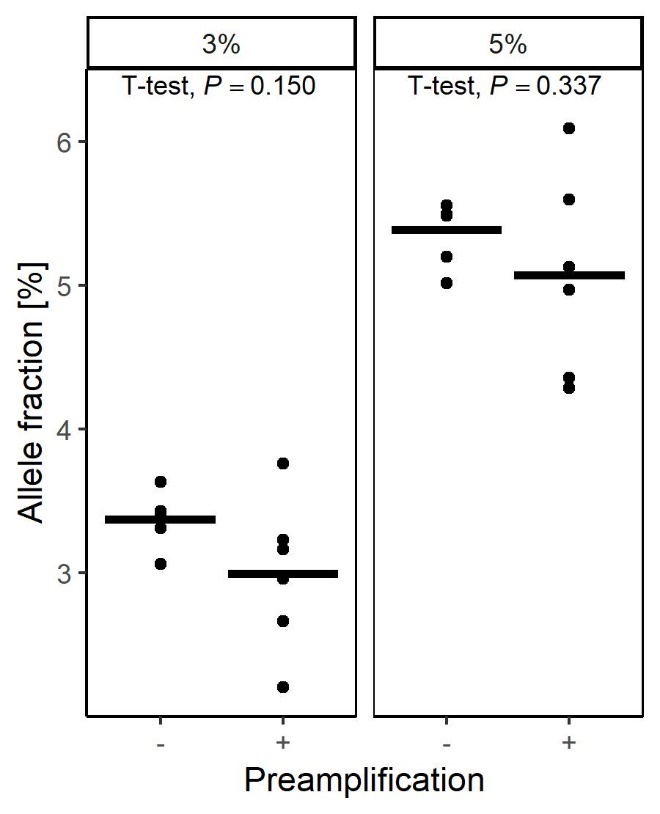

**Supplementary Figure 3: Targeted multiplex PCR preamplification does not change the mean allele fractions of six SNP loci in genomic DNA mixtures.** The genomic DNA mixtures were prepared by adding 3 or 5% (w/w) of genomic DNA of one genotype to genomic DNA of the different genotype. Dots represent mean allele fractions of the triplicate measurements with individual SNP assays and lines represent overall means. Shapiro-Wilk test was used to confirm normal distribution of the data and T-test was used for comparison of mean allele fractions without (-) and with (+) preamplification.

Next, we tested whether possible left-over chemicals in cfDNA extraction eluates affect PCR preamplification, that is inhibit or change SNP allele fractions (Suppl. Figure 4). In these experiments, we used mixtures of 161 bp long double-stranded gene fragments (gBlocks; Integrated DNA Technologies), with sequences corresponding to genomic regions that included the six studied SNPs. Specifically, PCR preamplification reactions of gBlocks as DNA templates for all six SNP amplicons (amplicons in 1:1 to 1:3 ratio between both possible alleles) were performed in the presence or absence of eluates of the mock extractions of cfDNA or evDNA, respectively (see Materials and Methods for DNA extraction protocols). All gBlock PCR preamplification reactions were prepared to mimic presence of whole volume of eluates in a final 50 or 100 µL reaction format, but were proportionally scaled down to a final volume of 20 µL. Preamplification of gBlocks showed 66 ± 10% (mean ± sd) efficiency, which was significantly decreased to 52 ± 10% and 48 ± 13% when whole eluates of the mock evDNA or cfDNA extractions were added to the preamplification reaction (mimicking 50 µL reaction format), respectively (*P* = 0.018 and *P* < 0.001; Suppl. Figure 4). When the eluates of the mock cfDNA or evDNA extractions were diluted two-times (100 µL reaction format), the efficiency returned to the levels observed for the preamplification of gBlocks in the absence of mock extractions (*P* = 0.268 and *P* = 0.999). Importantly, all tested allele fractions were comparable in gBlock samples with or without PCR preamplification, as determined by ddPCR (data not shown).

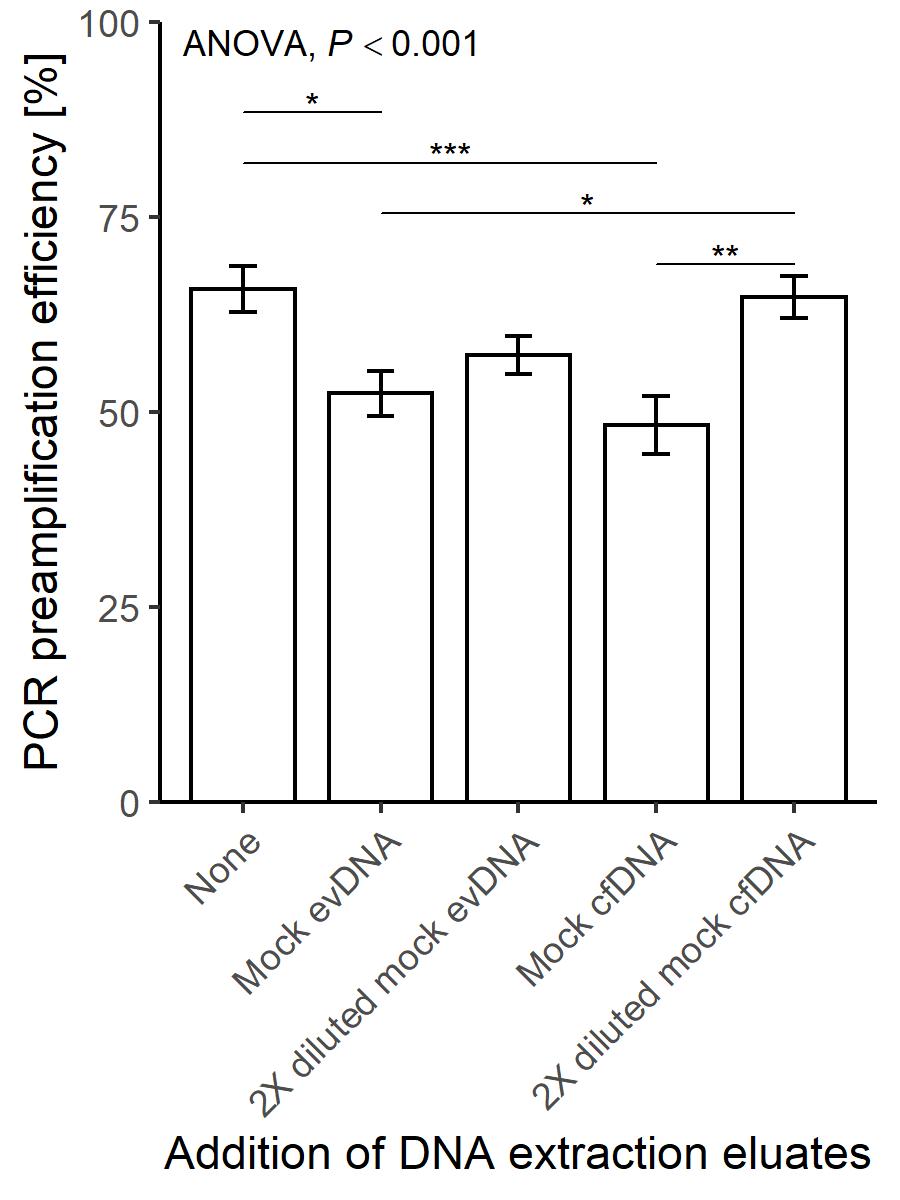

**Supplementary Figure 4: Evaluation of the effect of eluates of mock extractions of cfDNA or evDNA on the efficiency of PCR preamplification of gBlocks.** Mock eluates were added in such volumes that reflected presence of whole evDNA eluate (20 µL) in either a 50 µL (mock evDNA) or a 100 µL (2X diluted mock evDNA) PCR reaction; and whole cfDNA eluate (15 µL) in either a 50 µL (mock cfDNA) or a 100 µL (2X diluted mock cfDNA) PCR reaction. Mean PCR preamplification efficiency of alleles of 6 SNP assays and standard error of mean are presented. PCR preamplification efficiency is defined as the ratio between observed copy number to theoretically expected copy number after preamplification. Shapiro-Wilk test was used to confirm normal distribution of the data, ANOVA was used for comparison of PCR preamplification efficiencies and Tukey’s HSD test was used for multiple pairwise comparisons (* P < 0.050, ** P < 0.010, *** P < 0.001).

Finally, we analyzed allele fractions in one of the urine cfDNA samples without preamplification and after 9 and 11 cycles of preamplification. Based on donor and recipient genotyping, the cfDNA sample was heterozygous at five SNP loci. 5.5 ng cfDNA was used in each preamplification reaction, and the same amount was also directly used in ddPCR reactions for samples in the absence of preamplification. ddPCR was performed for all six SNP amplicons, in triplicates, as previously described. Preamplification efficiency was 68 ± 8% (mean ± sd) in reaction with 9 cycles and 65 ± 9% in reaction with 11 cycles (data not shown), which is in line with preamplification efficiency calculated for gBlocks. Importantly, mean allele fractions did not differ significantly (*P* = 0.700) in the absence or presence of preamplification, and were close to the expected 50% (Suppl. Figure 5).

Additionally, RPPH1/RPP30 ratio was determined for PCR-preamplified urine cfDNA, and compared to the determined ratio in samples without preamplification. Although RPPH1/RPP30 ratio was similar in the absence or presence of preamplification, certain variability in the range of expected coefficient of variation was observed (0.35 without preamplification, 0.40 after 9 cycles and 0.31 after 11 cycles).

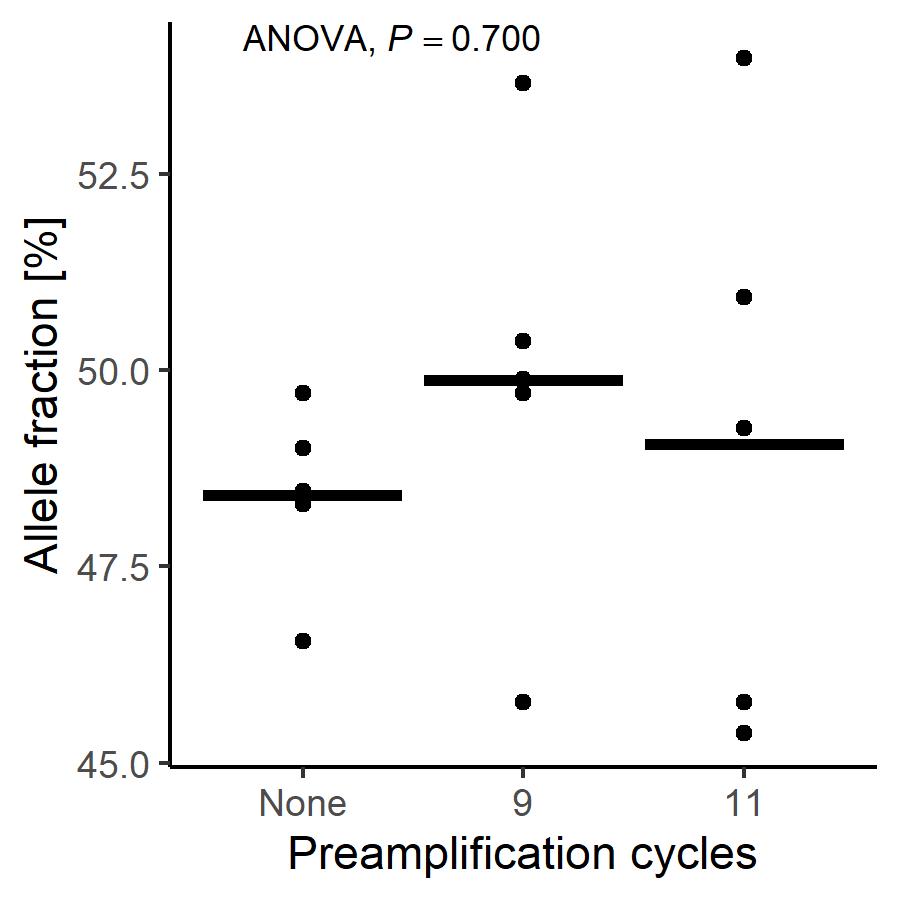

**Supplementary Figure 5: PCR preamplification does not change SNP allele fractions in a heterozygous urine cfDNA sample.** Dots represent mean allele fractions of the triplicate measurements with individual SNP assays and lines represent overall means. Shapiro-Wilk test was used to confirm normal distribution of the data and ANOVA was used for comparison of mean allele fractions in samples without preamplification and after 9 and 11 cycles of preamplification.

Based on all above results, we decided to proceed with targeted multiplex PCR preamplification of two-fold diluted total cfDNA and evDNA isolates in a 100 µL reaction format. We concluded that preamplifications preserves allele fractions and can be reliably used to determine the ddDNA fractions and ddDNA copies for extracted cfDNA and evDNA. In the case of RPPH1/RPP30 ratio (defined as integrity index), about 15% measurement error should be considered.

**S4 Uromodulin filament is non-specifically labelled with secondary IgG antibodies in immunogold TEM**

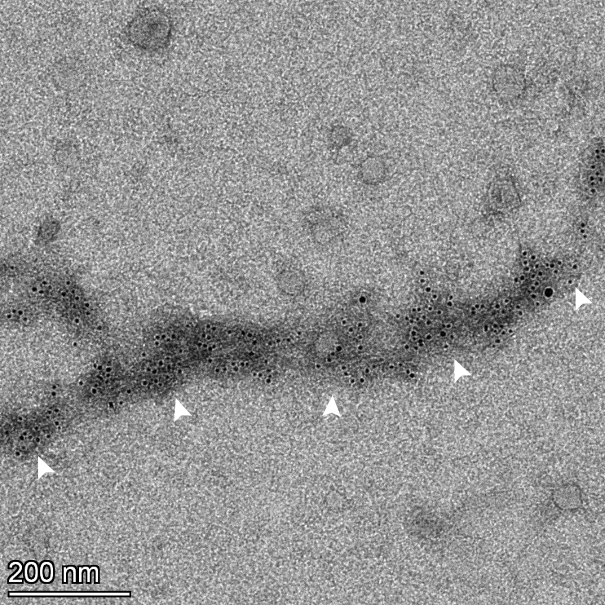

**Supplementary figure 6:** N**on-specific labelling of uromodulin filament in immunogold TEM.** TEM micrograph after immunogold labelling. Micrograph of uEV sample labelled with IgG antibodies conjugated with gold particles (6 nm), in the absence of antibodies against dsDNA. White arrows mark colloidal gold particles bound to uromodulin filaments.

**S5 Association of uEV, evDNA and cfDNA characteristics with kidney allograft recipients’ clinical characteristics**

**Supplementary Table 2**: Association of uEV characteristics with clinical parameters of the kidney allograft recipients’ continuous variables.

| **Variables** | **uEV concentration* [**× **10^10^/mmol]**  Spearman's rho  *P* | **uEV mean size [nm]**  Spearman's rho  *P* | **uEV mode size [nm]**  Spearman's rho  *P* | **uEV median size [nm]**  Spearman's rho  *P* |
| --- | --- | --- | --- | --- |
| Patient age | -0.147 | 0.106 | 0.081 | 0.147 |
|  | 0.386 | 0.515 | 0.619 | 0.367 |
| Tx to Bx [days] | 0.113 | 0.229 | 0.205 | 0.246 |
|  | 0.506 | 0.156 | 0.204 | 0.126 |
| HLA-mm | -0.040 | 0.167 | 0.031 | 0.194 |
|  | 0.815 | 0.304 | 0.850 | 0.231 |
| S-creatinine [µmol/L] | -0.167 | -0.047 | -0.037 | -0.046 |
|  | 0.324 | 0.774 | 0.822 | 0.777 |
| eGFR [mL/min/ 1.73m^2^] | 0.224 | -0.091 | -0.042 | -0.085 |
|  | 0.182 | 0.575 | 0.798 | 0.603 |
| S-CRP [mg/L] | -0.090 | -0.039 | 0.006 | 0.016 |
|  | 0.597 | 0.812 | 0.972 | 0.924 |

*uEV concentrations were normalized to urinary creatinine [mmol/L]. Abbreviations; uEV: urinary extracellular vesicle(s), Tx: transplantation, Bx: biopsy, HLA-mm: human leukocyte antigen mismatch, S-creatinine: serum creatinine, eGFR: estimated glomerular filtration rate, S-CRP: serum C-reactive protein.

**Supplementary Table 3**: Association of uEV characteristics with kidney allograft recipients’ clinical and histological characteristics (categorical variables).

| **Variables** | | **uEV concentration ****  **[**× **10^10^/mmol]**  median (25-75%) | **uEV mean size**  **[nm]**  median (25-75%) | **uEV mode size**  **[nm]**  median (25-75%) | **uEV median size**  **[nm]**  median (25-75%) |
| --- | --- | --- | --- | --- | --- |
| Sex | Male | 8.33 (3.65-13.89) [2] | 164.2 (144.8-176.8) | 121.9 (108.4-140.4) | 148.9 (129.7-163.2) |
|  | Female | 8.61 (2.79-32.79) [1] | 182.2 (174.2-197.6) | 141.4 (124.7-174.1) | 170.8 (156.2-190.0) |
|  | *P*-value | 0.894 | **0.004** | **0.043** | **0.009** |
| DGF | No | 8.53 (3.39-16.31) [3] | 172.3 (157.9-185.1) | 126.3 (112.9-144.8) | 154.9 (139.7-172.2) |
|  | Yes | 6.09 (3.90-14.73) | 149.6 (145.6-170.5) | 116.5 (102.8-142.5) | 134.0 (129.3-161.3) |
|  | *P*-value | 0.903 | 0.075 | 0.262 | 0.092 |
| ECD | No | 8.54 (2.92-23.02) [1] | 175.3 (148.8-185.0) | 127.8 (110.4-154.2) | 161.5 (133.7-174.0) |
|  | Yes | 8.20 (4.93-12.16) [1] | 165.6 (147.8-173.6) | 121. (113.2-139.6) | 151.6 (133.8-157.6) |
|  | *P*-value | 0.935 | 0.357 | 0.460 | 0.497 |
| Bx | Protocol | 6.77 (3.39-16.53) [1] | 166.5 (145.6-179.4) | 118.3 (110.3-141.8) | 149 (131.5-165.2) |
|  | For-cause | 8.61 (4.51-14.64) [2] | 174.5 (153.7-184.9) | 127.8 (115.7-153.0) | 161.5 (139.7-173.9) |
|  | *P*-value | 0.517 | 0.452 | 0.436 | 0.376 |
| t | Score 0 | 8.53 (3.78-16.53) [1] | 170 (148.4-179.7) | 126 (114-144.4) | 153.3 (133.8-170.8) |
|  | Score > 0 | 8.20 (3.12-14.33) [2] | 158.5 (142.5-186.9) | 122.9 (102.9-149.9) | 139.7 (127.6-172.2) |
|  | *P*-value | 0.651 | 0.807 | 0.577 | 0.702 |
| i | Score 0 | 7.78 (3.50-12.56) [1] | 169.8 (148.4-178.5) | 125.5 (112.7-142.3) | 153 (133.8-165.7) |
|  | Score > 0 | 11.14 (8.40-18.58) [2] | 184.9 (142.5-198.5) | 133.4 (109.6-155.5) | 171.5 (127.6-186.6) |
|  | *P*-value | 0.213 | 0.601 | 0.485 | 0.601 |
| ti | Score 0 | 8.60 (3.40-16.92) | 168.1 (144.8-174.7) | 124.8 (109.1-143.1) | 152.3 (129.7-160.7) |
|  | Score > 0 | 8.60 (3.60-17.52) [3] | 174.2 (157.3-185.5) | 128.7 (114.1-153.5) | 158.1 (139.7-173.9) |
|  | *P*-value | 1.000 | 0.217 | 0.593 | 0.276 |
| *g+ptc | Score 0 | 8.15 (3.62-12.56) [1] | 166.5 (147.2-178.8) | 125.5 (111.1-142.7) | 151.6 (132.5-168.0) |
|  | Score > 0 | 9.75 (5.68-17.66) [2] | 179.4 (149.6-186.9) | 133.4 (118.3-154.9) | 167.6 (134.0-173.9) |
|  | *P*-value | 0.620 | 0.442 | 0.485 | 0.485 |
| i-IFTA | Score 0 | 8.93 (3.10-16.92) | 165.6 (144.8-171.7) | 121.2 (112.6-143.7) | 150.9 (130.3-155.4) |
|  | Score > 0 | 8.61 (3.87-17.66) [2] | 173.9 (158.5-185.1) | 127.8 (115.7-153) | 158.1 (139.7-173.9) |
|  | *P*-value | 0.821 | 0.143 | 0.451 | 0.192 |

[No. of missing data]; score > 0: combined Banff Lesion Scores 1, 2 and 3; ** uEV concentration values were normalized to urinary creatinine [mmol/L]. Abbreviations: uEV: urinary extracellular vesicle(s), DGF: delayed graft function, ECD: expanded criteria donor, Bx: biopsy, Protocol: surveillance biopsy, t: tubulitis, i: interstitial inflammation, ti: total inflammation, *g+ptc: patients with glomerulitis (g) and peritubular capillaritis (ptc) combined, i-IFTA: inflammation in areas of fibrosis.

**Supplementary Table 4**: Association of uEV, evDNA and cfDNA characteristics with kidney allograft rejection phenotype.

|  |  | **Parameters/Variables** | **TCMR (N = 6)** | **ABMR (N = 4)** | ***P*-value** |
| --- | --- | --- | --- | --- | --- |
| **uEV characteristics** | **U-creatinine normalized** | **uEV oncentration**  **[**× **10^10^/mmol**  **U-creatinine]**  median (25-75%) | 8.20  (3.12-27.92) [1] | 9.75  (9.18-13.63) [1] | 0.571 |
|  | **Absolute values** | **uEV mean size [nm]**  median (25-75%) | 156.1  (142.4-191.8) | 185.9  (173.8-195.6) | 0.257 |
|  |  | **uEV mode size [nm]**  median (25-75%) | 116.2  (102.7-162.5) | 144.1  (126.1-155.3) | 0.257 |
|  |  | **uEV median size [nm]**  median (25-75%) | 139.6  (127.4-181.4) | 172.7  (157.6-183.4) | 0.257 |
| **evDNA** | **Expressed as fractions** | **evDNA integrity index [RPPH1/RPP30]**  median (25-75%) | 0.33  (0.26-0.51) | 0.43  (0.3-0.54) | 0.610 |
|  |  | **dd-evDNA fraction [%]**  median (25-75%) | 65.54  (41.29-82.74) | 32.72  (7.83-77.4) | 0.352 |
|  | **U-creatinine**  **normalized** | **evDNA yield [ng]**  median (25-75%) | 29.6  (16.8-174.6) [1] | 122.5  (80-208.6) [1] | 0.393 |
|  |  | **evDNA copy number [copies/mmol]**  median (25-75%) | 1000.8  (107.25-6167.25) [1] | 20881.1  (12970.7-41611.35) [1] | 0.071 |
|  |  | **dd-evDNA copy number [copies/mmol]**  **median (25-75%)** | 680.9  (62.4-1180.85) [1] | 3044.3  (2808.95-3260.8) [1] | **0.036** |
| **cfDNA** | **Expressed as fractions** | **cfDNA integrity index**  **[RPPH1/RPP30]**  median (25-75%) | 0.21  (0.17-0.24) | 0.4  (0.24-0.43) | 0.114 |
|  |  | **dd-cfDNA fraction [%]**  median (25-75%) | 64.81  (41.01-84.8) | 34.26  (8.66-58.01) | 0.171 |
|  | **U-creatinine**  **normalized** | **cfDNA yield [ng]**  median (25-75%) | 805.3  (307.6-3314) [1] | 4153.8  (3466.35-6611.8) [1] | 0.143 |
|  |  | **cfDNA copy number [copies/mmol]**  median (25-75%) | 59993.3  (20172.5-155209.45) [1] | 438517.7  (245836.55-1307061.3) [1] | 0.250 |
|  |  | **dd-cfDNA copy number**  **[copies/mmol]**  median (25-75%) | 19031  (10855.95-64952.8) [1] | 68083  (48124-103358) [1] | 0.250 |

[No. of missing data]; interquartile range was determined using weighted averages if more than 3 samples were included in the group and using Tukey's hinges if 3 samples were included in the group. Abbreviations; uEV: urinary extracellular vesicle(s), evDNA: extracellular vesicle-bound DNA, dd-evDNA: donor-derived extracellular vesicle-bound DNA, cfDNA: cell-free DNA, dd-cfDNA: donor-derived cell-free DNA, TCRM: T-cell mediated rejection, ABMR: antibody mediated rejection.

**Supplementary Table 5**: Association of evDNA characteristics with kidney allograft recipients’ clinical characteristics (continuous variables).

| **Variables** |  | | **U-creatinine normalized** | | |
| --- | --- | --- | --- | --- | --- |
|  | **evDNA**  **integrity index**  Spearman's rho  *P* | **dd-evDNA fraction [%]**  Spearman's rho  *P* | **evDNA yield [ng]**  Spearman's rho  *P* | **evDNA copy number**  **[copies/mmol]**  Spearman's rho  *P* | **dd-evDNA copy number**  **[copies/mmol]**  Spearman's rho  *P* |
| Patient age | -0.100 | -0.118 | -0.134 | -0.056 | -0.006 |
|  | 0.579 | 0.512 | 0.431 | 0.757 | 0.973 |
| Tx to Bx [days] | -0.003 | -0.080 | 0.120 | -0.047 | -0.088 |
|  | 0.987 | 0.660 | 0.481 | 0.793 | 0.644 |
| HLA-mm | 0.015 | -0.025 | 0.074 | -0.137 | -0.084 |
|  | 0.936 | 0.891 | 0.665 | 0.447 | 0.657 |
| S-creatinine [µmol/L] | 0.210 | -0.101 | -0.066 | -0.044 | -0.075 |
|  | 0.240 | 0.577 | 0.699 | 0.809 | 0.695 |
| eGFR [mL/min/ 1.73m^2^] | -0.307 | 0.284 | 0.035 | -0.013 | 0.092 |
|  | 0.082 | 0.109 | 0.838 | 0.941 | 0.628 |
| S-CRP [mg/L] | 0.023 | 0.105 | -0.119 | -0.335 | -0.199 |
|  | 0.899 | 0.562 | 0.484 | 0.057 | 0.293 |

Abbreviations; evDNA: extracellular vesicle-bound DNA, dd-evDNA: donor-derived extracellular vesicle-bound DNA, Tx: transplantation, Bx: biopsy, HLA-mm: human leukocyte antigen mismatch, S-creatinine: serum creatinine, eGFR: estimated glomerular filtration rate, S-CRP: serum C-reactive protein.

**Supplementary Table 6**: Association of cfDNA characteristics with kidney allograft recipients’ clinical characteristics (continuous variables).

| **Variables** |  | | **U-creatinine normalized** | | |
| --- | --- | --- | --- | --- | --- |
|  | **cfDNA**  **integrity index**  Spearman's rho  *P* | **dd-cfDNA fraction [%]** Spearman's rho  *P* | **cfDNA yield [ng]** Spearman's rho  *P* | **cfDNA copy number**  **[copies/mmol]**  Spearman's rho  *P* | **dd-cfDNA copy number**  **[copies/mmol]**  Spearman's rho  *P* |
| Patient age | -0.039 | -0.018 | -0.079 | -0.155 | -0.022 |
|  | 0.813 | 0.912 | 0.643 | 0.368 | 0.900 |
| Tx to Bx [days] | 0.030 | -0.127 | 0.009 | -0.022 | -0.125 |
|  | 0.854 | 0.434 | 0.958 | 0.898 | 0.467 |
| HLA-mm | 0.112 | -0.172 | 0.102 | 0.088 | 0.041 |
|  | 0.497 | 0.289 | 0.549 | 0.610 | 0.811 |
| S-creatinine [µmol/L] | -0.035 | -0.021 | -0.109 | -0.161 | -0.158 |
|  | 0.833 | 0.895 | 0.519 | 0.347 | 0.356 |
| eGFR [mL/min/ 1.73m^2^] | -0.115 | 0.226 | -0.006 | 0.059 | 0.172 |
|  | 0.485 | 0.161 | 0.972 | 0.732 | 0.316 |
| S-CRP [mg/L] | 0.069 | 0.107 | -0.309 | -0.287 | -0.280 |
|  | 0.674 | 0.511 | 0.062 | 0.090 | 0.098 |

Abbreviations; cfDNA: cell-free DNA, dd-cfDNA: donor-derived cell-free DNA, Tx: transplantation, Bx: biopsy, HLA-mm: human leukocyte antigen mismatch, S-creatinine: serum creatinine, eGFR: estimated glomerular filtration rate, S-CRP: serum C-reactive protein.
