## Supplementary material for "Extracellular vesicle-bound DNA in urine is indicative of kidney allograft injury": MISEV2018 Checklist

Numbers refer to sections listed in the Table of contents from: C. Théry and K.W.Witwer, et al, "Minimal Information for Studies of Extracellular Vesicles 2018 (MISEV2018): a posi- tion statement of the International Society for Extracellular Vesicles and update of the MISEV2014 guidelines", J Extracell Vesicles 2018;7:1535750.

O Mandatory O Mandatory if applicable O Encouraged

### 1-Nomenclature

Mandatory

X Generic term extracellular vesicle (EV): With demon- stration of extracellular (no intact cells) and vesicular nature per these characterization (Section 4) and function (Section 5) guidelines OR

O Generic term, e.g., extracellular particle (EP): no intact cells but MISEV guidelines not satisfied

Encouraged (choose one)

X Generic term extracellular vesicle (EV) + specification (size, density, other)

O Specific term for subcellular origin: e.g., ectosome, microparticle, microvesicle (from plasma membrane), exosome (from endosomes), with demonstration of the subcellular origin

O Other specific term: with definition of specific criteria

### 2-Collection and pre-processing

Tissue Culture Conditioned medium (CCM, Section 2-a) O General cell characterization (identity, passage, myco- plasma check…)

O Medium used before and during collection (additives, serum, other)

O exact protocol for depletion of EVs/EPs from addi- tives in collection medium

O Nature and size of culture vessels, and volume of medium during conditioning

O specific culture conditions (treatment, % O2, coating, polarization…) before and during collection

O Number of cells/ml or /surface area and % of live/ dead cells at time of collection (or at time of seeding with estimation at time of collection)

O Frequency and interval of CM harvest

Biofluids or Tissues (Sections 2-b and -c)

X Donor status if available (age, sex, food/water intake, collection time, disease, medication, other)

X Volume of biofluid or volume/mass of tissue sample collected per donor

O Total volume/mass used for EV isolation (if pooled from several donors)

X All known collection conditions, including additives, at time of collection

X Pre-treatment to separate major fluid-specific con- taminants before EV isolation

X Temperature and time of biofluid/tissue handling before and during pre-treatment

O For cultured tissue explants: volume, nature of med- ium and time of culture before collecting conditioned medium

O For direct tissue EV extraction: treatment of tissue to release vesicles without disrupting cells

Storage and recovery (Section 2-d)

X Storage and recovery (e.g., thawing) of CCM, bio- fluid, or tissue before EV isolation (storage tempera- ture, vessel, time; method of thawing or other sample preparation)

X Storage and recovery of EVs after isolation (tempera- ture, vessel, time, additive(s)…)

### 3-EV separation and concentration

Experimental details of the method

O Centrifugation: reference number of tube(s), rotor(s), adjusted k factor(s) of each centrifugation step (= time+ speed+ rotor, volume/density of centrifugation condi- tions), temperature, brake settings

O Density gradient: nature of matrix, method of gen- erating gradient, reference (and size) of tubes, bottom- up (sample at bottom, high density) or top-bottom (sample on top, low density), centrifugation speed and time (with brake specified), method and volume of fraction recovery

X Chromatography: matrix (nature, pore size,…), loaded sample volume, fraction volume, number

O Precipitation: reference of polymer, ratio vol/vol or weight/vol polymer/fluid, time/temperature of incuba- tion, time/speed/temperature of centrifugation

O Filtration: reference of filter type (=nature of mem- brane, pore size…), time and speed of centrifugation, volume before/after (in case of concentration)

O Antibody-based : reference of antibodies, mass Ab/ amount of EVs, nature of Ab carrier (bead, surface) and amount of Ab/carrier surface

O Other…: all necessary details to allow replication

O Additional step(s) to concentrate, if any

O Additional step(s) to wash matrix and/or sample, if any

Specify category of the chosen EV separation/concentra- tion method (Table 1):

O High recovery, low specificity = mixed EVs and non- EV components OR

X Intermediate recovery, intermediate specificity = mixed EVs with limited non-EV components OR

O Low recovery, high specificity = subtype(s) of EVs with as little non-EV as possible OR

O High recovery, high specificity = subtype(s) of EVs with as little non-EV as possible

### 4-EV characterization

Quantification (Table 2a, Section 4-a)

X Volume of fluid, and/or cell number, and/or tissue mass used to isolate EVs

O Global quantification by at least 2 methods: protein amount, particle number, lipid amount, expressed per volume of initial fluid or number of producing cells/ mass of tissue

O Ratio of the 2 quantification figures

Global characterization (Section 4-b, Table 3) *

X Transmembrane or GPI-anchored protein localized in cells at plasma membrane or endosomes

X Cytosolic protein with membrane-binding or - association capacity

X Assessment of presence/absence of expected contaminants (At least one each of the three categories above)

O Presence of proteins associated with compartments other than plasma membrane or endosomes

O Presence of soluble secreted proteins and their likely transmembrane ligands

O Topology of the relevant functional components (Section 4-d)

Single EV characterization (Section 4-c)

O Images of single EVs by wide-field and close-up: e.g. electron microscopy, scanning probe microscopy, super-resolution fluorescence microscopy

X Non-image-based method analysing large numbers of single EVs: NTA, TRPS, FCS, high-resolution flow cytometry, multi-angle light-scattering, Raman spectroscopy, etc.

5-Functional studies

O Dose-response assessment

O Negative control = nonconditioned medium, bio-fluid/tissue from control donors, as applicable

O Quantitative comparison of functional activity of total fluid, vs EV-depleted fluid, vs EVs (after high recovery/low specificity separation)

O Quantitative comparison of functional activity of EVs vs other EPs/fractions after low recovery/high spe- cificity separation

O Quantitative comparison of activity of EV subtypes (if subtype-specific function claimed)

O Extent of functional activity in the absence of contact between EV donor and EV recipient

6-Reporting

X Submission of methodologic details to EV-TRACK (evtrack.org) with EV-TRACK number provided (strongly encouraged)

O Submission of data (proteomic, sequencing, other) to relevant public, curated databases or open-access repositories

O Data submission to EV-specific databases (e.g., EVpedia, Vesiclepedia, exRNA atlas)

O Temper EV-specific claims when MISEV requirements cannot be entirely satisfied (Section 6-b)

* Global characterization relevant information reported in:
Sedej, I., Žnidarič, M. T., Dolžan, V., Lenassi, M., & Arnol, M. (2021).
Optimization of isolation protocol and characterization of urinary extracellular vesicles as biomarkers of kidney allograft injury. Clinical Nephrology, 96(Suppl 1), 107–113. https://doi.org/10.5414/CNP96S19
