## Supplementary material for "Extracellular vesicle-bound DNA in urine is indicative of kidney allograft injury": REMARK for EQUATOR network Checklist

| **Item to be reported** | | **Page no.** |
| --- | --- | --- |
| **INTRODUCTION** | |  |
| 1 | State the marker examined, the study objectives, and any pre-specified hypotheses. | 4 |
| **MATERIALS AND METHODS** | |  |
| *Patients* | |  |
| 2 | Describe the characteristics (e.g., disease stage or co-morbidities) of the study patients, including their source and inclusion and exclusion criteria. | 5 |
| 3 | Describe treatments received and how chosen (e.g., randomized or rule-based). | 5 |
| *Specimen characteristics* | |  |
| 4 | Describe type of biological material used (including control samples) and methods of preservation and storage. | 5, 6 |
| *Assay methods* | |  |
| 5 | Specify the assay method used and provide (or reference) a detailed protocol, including specific reagents or kits used, quality control procedures, reproducibility assessments, quantitation methods, and scoring and reporting protocols. Specify whether and how assays were performed blinded to the study endpoint. | 6-10 |
| *Study design* | |  |
| 6 | State the method of case selection, including whether prospective or retrospective and whether stratification or matching (e.g., by stage of disease or age) was used. Specify the time period from which cases were taken, the end of the follow-up period, and the median follow-up time. | 5 |
| 7 | Precisely define all clinical endpoints examined. | 5 |
| 8 | List all candidate variables initially examined or considered for inclusion in models. | 5-9 |
| 9 | Give rationale for sample size; if the study was designed to detect a specified effect size, give the target power and effect size. | 5 |
| *Statistical analysis methods* | |  |
| 10 | Specify all statistical methods, including details of any variable selection procedures and other model-building issues, how model assumptions were verified, and how missing data were handled. | 10 |
| 11 | Clarify how marker values were handled in the analyses; if relevant, describe methods used for cutpoint determination. | 9-12 |
| **RESULTS** | |  |
| *Data* | |  |
| 12 | Describe the flow of patients through the study, including the number of patients included in each stage of the analysis (a diagram may be helpful) and reasons for dropout. Specifically, both overall and for each subgroup extensively examined report the numbers of patients and the number of events. | 11 |
| 13 | Report distributions of basic demographic characteristics (at least age and sex), standard (disease-specific) prognostic variables, and tumor marker, including numbers of missing values. | 27 |
| *Analysis and presentation* | |  |
| 14 | Show the relation of the marker to standard prognostic variables. | n.a. |
| 15 | Present univariable analyses showing the relation between the marker and outcome, with the estimated effect (e.g., hazard ratio and survival probability). Preferably provide similar analyses for all other variables being analyzed. For the effect of a tumor marker on a time-to-event outcome, a Kaplan-Meier plot is recommended. | 11-15 |
| 16 | For key multivariable analyses, report estimated effects (e.g., hazard ratio) with confidence intervals for the marker and, at least for the final model, all other variables in the model. | n.a. |
| 17 | Among reported results, provide estimated effects with confidence intervals from an analysis in which the marker and standard prognostic variables are included, regardless of their statistical significance. | n.a. |
| 18 | If done, report results of further investigations, such as checking assumptions, sensitivity analyses, and internal validation. | n.a. |
| **DISCUSSION** | |  |
| 19 | Interpret the results in the context of the pre-specified hypotheses and other relevant studies; include a discussion of limitations of the study. | 15-19 |
| 20 | Discuss implications for future research and clinical value. | 19 |
